# Modeling the Dynamics of COVID-19: Model With Patients With underlying conditions

**DOI:** 10.1101/2025.08.14.25332881

**Authors:** Geoffrey Magak, Josephine Kagunda, Marilyn Rono

## Abstract

**Introduction:** Coronavirus disease (COVID-19) is an infectious disease caused by the SARS-CoV-2 virus. The virus can spread from an infected person’s mouth or nose in small liquid particles when they cough, sneeze, speak, sing or breathe. These particles range from larger respiratory droplets to smaller aerosols.

**Objective:** The study aims to formulate a deterministic model to understand the dynamics of SARs Corona Virus infection among the Kenyan population with a vital interest in two proportions; people with other chronic illnesses and those who do not have other chronic illnesses.

**Methodology:** We formulate, study and analyze a deterministic model of COVID-19 transmission dynamics with eight compartments. Here, we use a system of non-linear ordinary differential equations with optimal control and three therapeutic measures, which include vaccination of the Susceptible proportion, treatment at the hospital, and homebased care mitigation and treatment. This system has disease-free and endemic equilibria points whose stability is investigated. The basic reproduction number, *R*_0_, is calculated using the next-generation matrix. The disease-free equilibrium, *DFE*_*q*_ of the model is asymptotically stable if *R*_0_ < 1 and unstable if *R*_0_ > 1 whereas endemic equilibrium, *EE*_*q*_ of the model becomes asymptotically stable if *R*_0_ >1. Sensitivity analysis was performed on all parameters to determine their impact on the transmission of COVID-19. Finally, the parameter estimation was conducted, and numerical simulation of the model using MATLAB and graphs were shown.

**Results:** A basic reproduction number,*R*_0_ = 1.35, indicating that COVID-19 will persist within the population. Results from the simulation suggest that low adherence to the measures put in place to curb the disease increases infection in the population. Hospitalization and home-based care programs show that an increased rate of hospitalization and care lowers infection.

## 1 Introduction

### 1.1 Background

#### 1.1.1 History of SARs Corona Virus

In December 2019, many patients reported to hospitals in Wuhan, China, with symptoms like pneumonia [1]. The number of patients admitted increased tremendously, and this made people worried. It was later determined that a virus had sprung up from Wuhan open market in Hubei province, China, which sold seafood and live animals like snakes and frogs[2]. This made Wuhan city the epicenter of this virus. The medics confirmed that the virus was hosted by animals found in that market.[1] -[2]. The virus was later identified as Severe Acute Respiratory Syndrome Coronavirus 2 (SARS-CoV-2) and was named Coronavirus disease 2019 (COVID-19) by the World Health Organization (WHO) [3].

#### 1.1.2 COVID-19 in Kenya

On 13 March 2020, Kenya had patient zero, who had traveled from England and thus made Nairobi city the epicenter.[7]. Through the Ministry of Health, the Kenyan government reminded the citizens that most infected persons experienced only mild illness and recovered easily. Still, the disease can be more severe in others, especially the elderly and persons with other chronic illnesses. In a few months, the virus had spread so first that it prompted the head of state with the Ministry of Health’s advice to bring into action a curfew and then a partial lockdown to curb the spread. The other preventive measures that were put in place to curb the spread were:

By the end of 2020, Kenya had recorded 1692 new cases within that week, and cumulative positive cases of 95, 843 [8] from a sample of 1,046,667, and 78,737 patients had recovered from the disease. However, it was unfortunate that 1670 people lost their lives.

### 1.2 Problem Statement

The COVID-19 virus began in 2019 in Wuhan, China, and has since spread worldwide. The virus spreads easily and can affect anyone who is exposed to it. This virus spreads faster according to adherence to laid-out mitigation measures.

In Kenya, the most affected groups are the elderly and those with other chronic illnesses, especially HIV/AIDS, heart disease, liver problems, kidney failure, and others. Older adults are more likely to get very sick from COVID-19. Getting very sick means that older adults with COVID-19 might need hospitalization, intensive care, or a ventilator to help them breathe, or they might even die[27]. The risk increases for people in their 50s and 60s, 70s, and 80s. People 85 and older are the most likely to get very sick. People with other chronic illnesses are more likely to get severely ill with COVID-19 [27]. A person’s risk of severe illness from COVID-19 increases as the number of underlying medical conditions they have increased.

To keep the spread rate safe and lower, we need to observe the mitigation procedure and processes to curb this spread.

Mathematical models have been used to study human problems, like solving problems associated with an outbreak of infectious diseases.Modeling infectious disease transmission is very important in the medical field.Glassy and Flasser (2008) noted that the models are applied to surveillance in addressing disease-control policy questions in making decisions about control and surveillance [10]. It informs the medics and the government on its rate of spread which is used in determining the peak and waves of the infected cases. The strategy is fundamental in understanding the spread and helps in planning for the best control strategies .[11] Since the outbreak of COVID-19, many scholars have used either deterministic or stochastic models. The deterministic model allows a researcher to do a calculation of a future event and it does not involve randomness. For instance, Ogana et al. did a SIRD model applied to COVID-19 dynamics and intervention strategies during the first wave in Kenya.[11] to understand how the intervention strategies influenced the spread.

To model infectious diseases like COVID-19, Ebola, and Malaria, various models are used to unearth the dynamics of their spread and peaks.[10, 12] The infectious disease models are categorized into compartmental or deterministic model[14],[18] and stochastic(random) model[15], [17]. The Monte Carlo simulation is an example of a stochastic model.

In deterministic models, there are compartments to the population that separate different population groups according to stages. This model is formulated by an ordinary differential equation (ODE) system based on the SEIRD framework.

Ogana, Juma, and Bulimo (2021) created a SIRD model to apply to the intervention strategies, and dynamics of COVID-19 during the first wave [11]. Their methods only applied to a single wave. Since we have had many waves, our study will incorporate them. Mathematical Modelling of COVID-19 Transmission in Kenya: A Model with Reinfection Transmission Mechanism by Wangari et al. l(2021) [14]. Mwalili et al. (2020) did an SEIR model for COVID-19 dynamics incorporating the environment, and social distancing [21]. Ki-mathi et al. (2020) on Age-structured model for COVID-19: Effectiveness of social distancing and contact reduction in Kenya [24]. Mbogo and Odhiambo (2021) did a study on the COVID-19 outbreak, social distancing and mass testing in Kenya-insights from a mathematical model [25].

From this study, we are going to make a comparison in the spread bettween people who have other chronic illnesses and those who do not. Vaccination will also be considered, in general, whether one has been fully or partially vaccinated.

### 1.3 Research Question

How does the infection load and progression of the virus differ among individuals with chronic illnesses in terms of infection rates, hospitalization, home-based care, and recovery outcomes compared to the general population?

### 1.4 Objectives

The study’s main objective is to formulate a deterministic model to understand the dynamics of the SARs Corona Virus infection rate among the Kenyan proportion of people with other chronic illnesses and those without.

The specific objectives are

a. Develop a compartmental model on COVID-19 transmission dynamics that will describe the transmission of this virus among patients with other chronic illnesses and the other proportion that do not have other chronic illnesses in Kenya.
b. Determine the basic reproduction number of the model.
c. Conduct mathematical analysis by obtaining numerical solutions of the deterministic model.

## 2 Model Description and Formulation

### 2.1 Model Description

Mathematical models play a key role in projecting how infectious diseases spread and whether there is a likelihood of becoming an epidemic. This helps in informing various health interventions.

The model uses statistical data to calculate parameters to monitor the effects and develop controls. For this, we modified the SEIR model to develop parameters that can inform the situation of COVID-19 infection among people with other chronic illnesses.

For this model, we show the transmission dynamics of people with and without chronic illnesses within the sub-populations of the exposed and infected.

The total population of people within the Kenyan boundaries at any time,t are considered to be susceptible, with a proportion of them vaccinated. When the susceptible population or the fully vaccinated comes into contact with the infected, they become exposed, E.

A proportion of the exposed get infected or rather test positive for COVID-19 and is presented by those Infected with other chronic illnesses *I*_*C*_ and infected with no other chronic illnesses *I*_*W*_.

The infected proportions are hospitalized, *H*_+_ or given the home-based care *H*_*B*_ program, where a caregiver monitors them. Home-based care infected proportion was introduced to capture the majority of the people who were nursed from home, taking natural remedies, and had very mild symptoms.

The hospitals and home-based care have patients who have been affected by the virus, and therefore quarantined to control the spread and manage their conditions.

The total population in Kenya thus at any time t is given by N(t)=S(t)+V(t)+E(t)+*I*_*C*_(*t*) +*I*_*W*_ (*t*) + *H*_+_(*t*) + *H*_*B*_(*t*)+R(t)

We assume that, there were no births, all COVID-19 cases were detected, all vaccinated individuals who caught COVID-19 and recovered developed herd immunity and all the exposed individuals with other chronic illnesses were infected due to low immunity.

### 2.2 Model Formulation

From the eight compartments, the susceptible population is the population of people within the Kenyan boundaries at the time,t. This proportion is recruited into the population by a constant immigration rate *λ* and a proportion of the people that have recovered and are prone to get reinfected, which is *υ*. Part of the susceptible are vaccinated at a rate of m, and the constant recruitment by constant immigration rate *λ*.

The Exposed population is a proportion of the susceptible or vaccinated population who have come into contact with the Infected. They get exposed to the virus infection at rate *σ* from the Susceptible and *ξσ* from the vaccinated, where *ξ* is the reduction rate of getting infected among individuals who have been partially or fully vaccinated. These people are identified through contact tracing from their interactions with the infected population. Some suspected they were exposed in Kenya made reports and were taken to quarantine facilities for monitoring.

The Exposed proportion gets infected after 7-14 days. This is the window period for the virus to show signs in an individual. A proportion of the exposed become infected with COVID-19 at rates *γ* for the people with other chronic illnesses and *π* and those who do not have other chronic illnesses. Some proportions of individuals who have been exposed and are not suffering from other chronic illnesses do not get infected and hence progress to the Susceptible population at a rate of *ω*.

The force of infection, 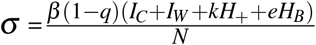 where *β* is the transmission rate, q is the population proportion which fails comply with COVID-19 guidelines, k is the percentage reduction of infections due to hospitalization, e is the percentage reduction of infections due to home-based care, and N is the total population.

Severe cases of the infected proportions with other chronic illnesses are hospitalized, *H*_+_, and mild cases are given the home-based care *H*_*B*_ program where a caregiver monitors them. The infected proportions are hospitalized if they exhibit very severe outcomes at a rate of *δ*. The other percentage of the infected opt or are advised to get home-based care at a rate of 1 \**δ* denoted as *δ*_1_.

Severe cases of the infected proportions with no other chronic illnesses are hospitalized, *H*_+_, and mild cases are given the home-based care *H*_*B*_ program where a caregiver monitors them. The infected proportions are hospitalized if they exhibit very severe outcomes at a rate of *θ*. The other percentage of the infected opt or are advised to get home-based care at a rate of 1 \**θ* denoted as *θ*_1_.

The recovery rate from the proportion that recovers from both the hospital and homebased care is *ψH*_+_ + *ϕH*_*B*_. Since some people did not develop immunity, they became susceptible after recovering. So the recovered R proportion becomes susceptible at a rate *υ*.

The natural death rate in every compartment is denoted by *µ*.COVID -19 related deaths are denoted by *τ*

We have created compartments of the population into eight S, V, E, *I*_*C*_, *I*_*W*_, *H*_+_, *H*_*B*_, and R.

### 2.3 Model Flow Diagram

### 2.4 Equations

From the model descriptions and the model flow diagram in Figure 1, the dynamics in the model system give the following equations:

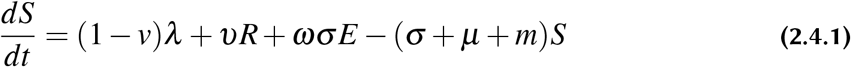

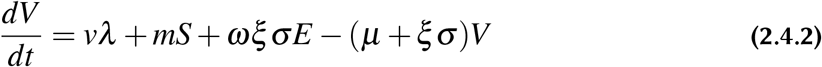

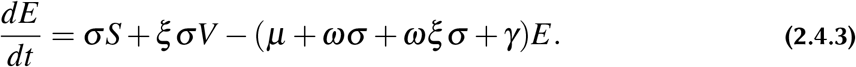

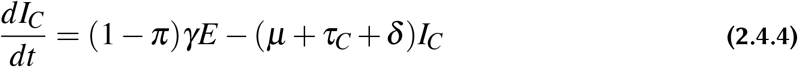

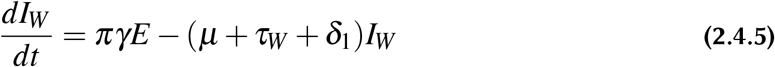

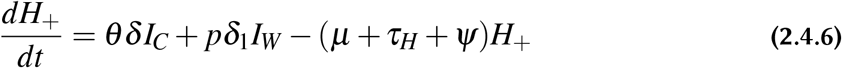

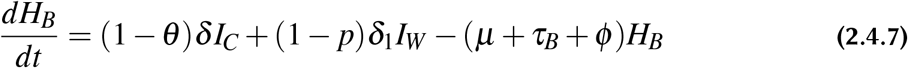

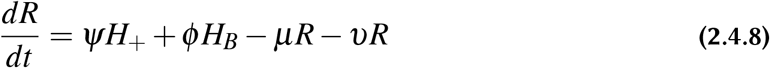

**Figure 1.**
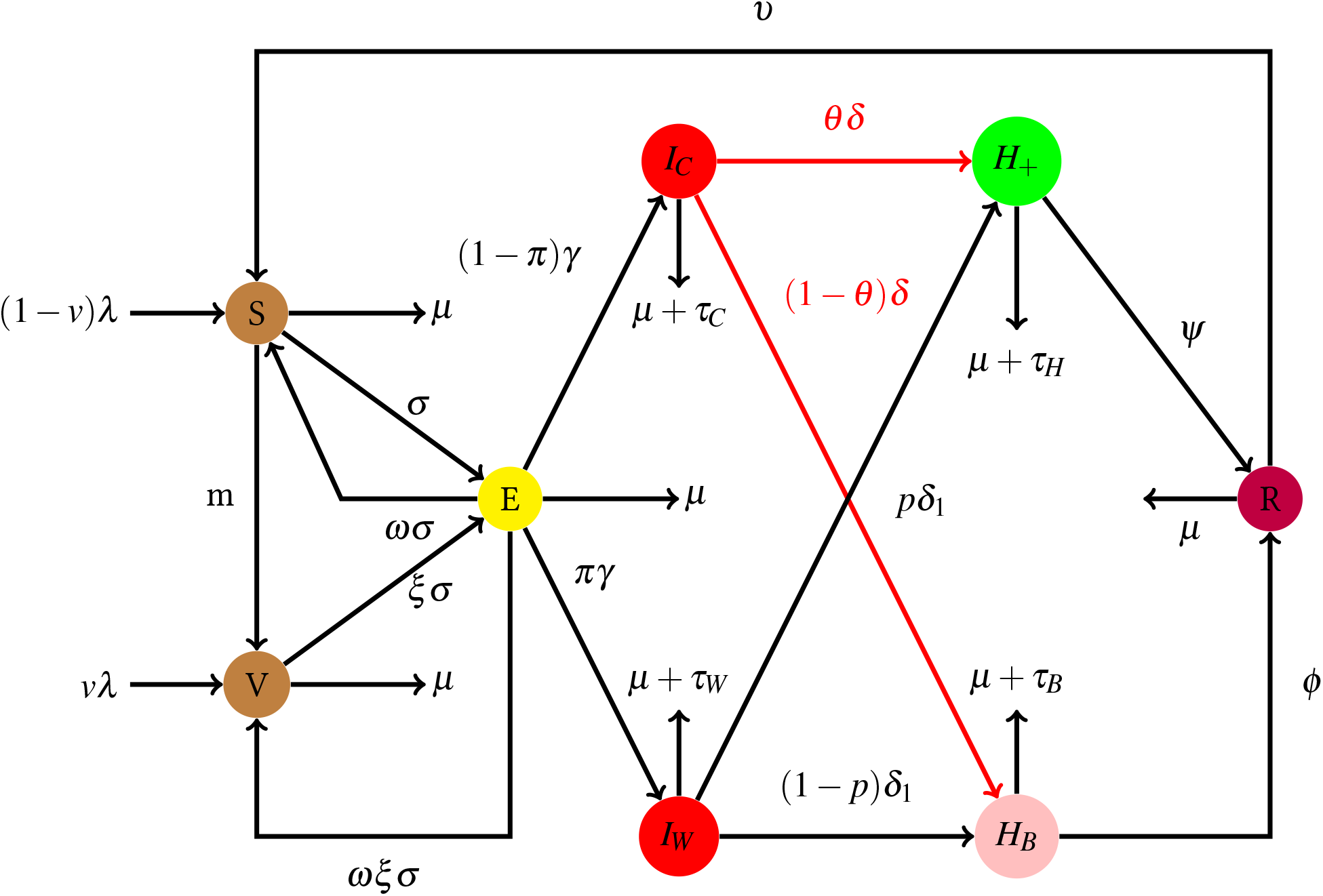
Model Flow Diagram for COVID-19 in Kenya.

## 3 Mathematical Analysis

### 3.1 Positivity and Boundedness

In this section, a proof that under non-negative conditions, the model system in figure 1 in chapter three has positive solutions in the compartments.

#### 3.1.1 Positivity

##### Theorem 1

Let S(t), V(t), E(t), *I*_*C*_(*t*), *I*_*W*_ (*t*), *H*_+_(*t*), *H*_*B*_(*t*), *R*(*t*) where t≥ 0 be the solution of equations 3.6.1 - 3.6.8 given the above condition, then *S*(0) ≥ 0, V(0)≥ 0, E(0)≥ 0, *I*_*C*_(0) ≥ 0, *I*_*W*_ (0) ≥ 0, *H*_+_(0) ≥ 0, *H*_*B*_(0) ≥ 0, R(0)≥ 0

The solutions of equtions 3.6.1 - 3.6.8 when t>0 that is S(t),V (t), E(t), *I*_*C*_(*t*), *I*_*W*_ (*t*), *H*_+_(*t*), *H*_*B*_(*t*), R(t) will be non negative numbers.

Proof

To prove this, we show that the equations 3.6.1 - 3.6.8 in the model are positive when t ≥0.

Let y(t)=S(t), V(t), E(t), *I*_*C*_(*t*), *I*_*W*_ (*t*), *H*_+_(*t*), *H*_*B*_(*t*), R(t) be the system under initial conditions *y*_0_=S(0), V(0), E(0), *I*_*C*_(0), *I*_*W*_ (0), *H*_+_(0), *H*_*B*_(0),

By the continuity of solution, for all the variables S(t), V(t), E(t), *I*_*C*_(*t*), *I*_*W*_ (*t*), *H*_+_(*t*), *H*_*B*_(*t*), R(t) that have the positive initial values at t=0, there exists an interval (0,*t*_*a*_) such that S(t), V(t), E(t), *I*_*C*_(*t*), *I*_*W*_ (*t*), *H*_+_(*t*), *H*_*B*_(*t*), R(t)≥ 0 for 0 *< t < t*_*a*_

For the susceptible,S

Lets consider 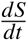

Suppose S=0, then

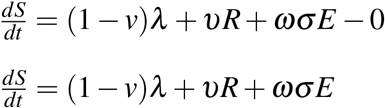

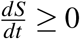 then the trajectory faces upwards when S=0.

If S=N, that is everyone is in Susceptible class then

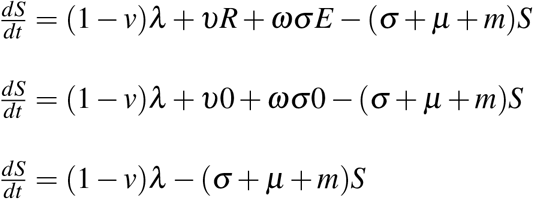

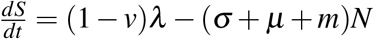, at any time, t, N is always greater than *λ*

So 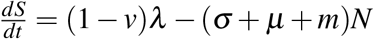 can also be expresed as

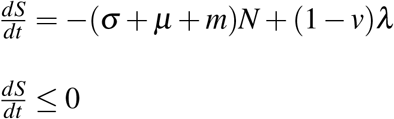

The trajectory faces downwards.

Hence, 0 ≥ *S*(*t*) ≤ *N* so the population solution for S will be positive for all t ≥ 0 This is true for other compartmets

#### 3.1.2 Boundedness

The model solutions are bounded in a set D

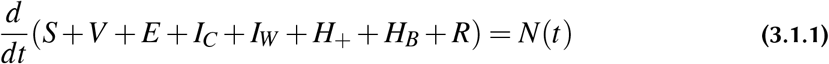

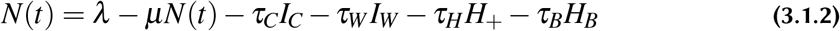

The death rates due to COVID-19 is a positive number hence it suffices to say that *τ*_*I*_, *τ*_*W*_, *τ*_*H*_, *τ*_*B*_>0. Therefore,

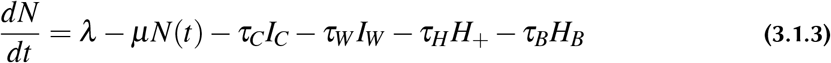

at the DFE.

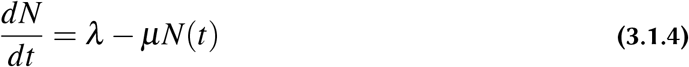

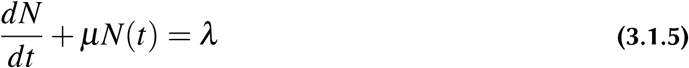

Using the time,t integrating factor

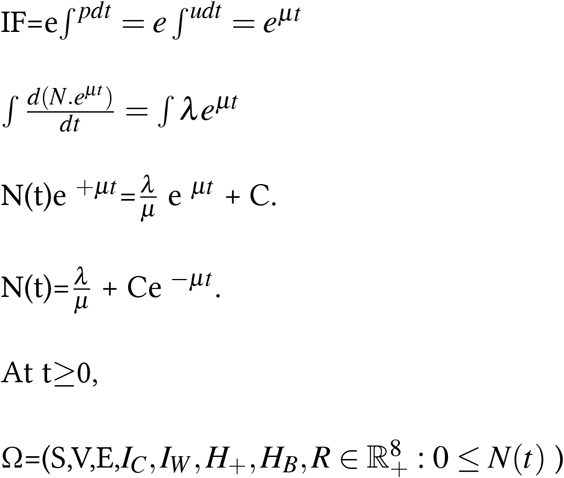

So it is bounded above.

### 3.2 Local existence and uniqueness

Let y = (*y*_*i*_)_*i*=1,2,…,8_ and f :ℝ_+_ x ℝ^8^ be continuous with respect to t, y and Lipschitz continuous. Let f(t, y) be non negative for all (t,y) ∈ ℝ_+_ X ℝ^8^ and *y*_*i*_ = 0. For every *y*_0_ ∈ T ℝ^8^ there exists a positive constant T such that 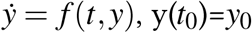 has a unique, positive and existing solution whose value lies in the interval [0, T) and in ℝ_+_^8^. If T<∞ then lim sup 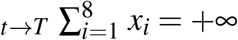

To prove this,

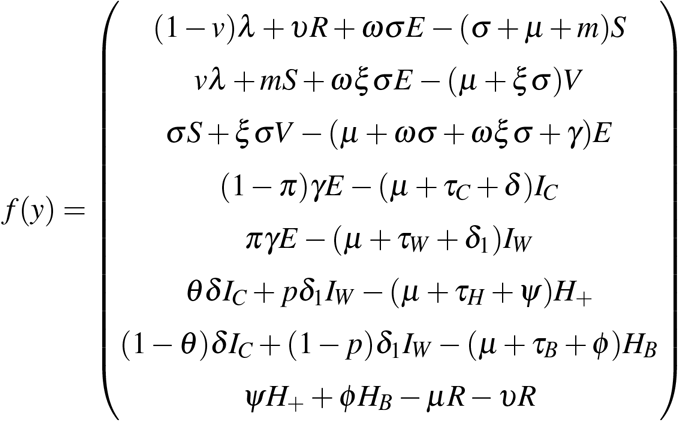

where y=[S,V,E,*I*_*C*_, *I*_*W*_, *H*_+_, *H*_*B*_, *R*]^*T*^. This means that f has a continuous partial derivative with respect to each state variables in 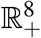 and are all bounded hence f is locally Lipschitz in 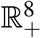.

### 3.3 Disease Free Equilibrium

The disease-free equilibrium point is a steady-state solution point where there are no new COVID-19 infections. This means that the infected with other chronic illnesses,*I*_*C*_, and those with no other chronic illnesses, *I*_*W*_, are zero, and the entire community has only the susceptible compartment.

In this scenario, the Exposed with chronic illnesses, *I*_*C*_, Exposed with no other chronic illnesses, *I*_*W*_, hospitalized, *H*_+_, home-based care patients, *H*_*B*_, and recovered, R are all zero.

Therefore, we use zero to get the disease-free equilibrium and set it on every equation.

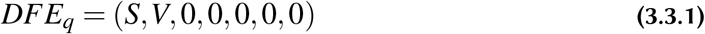

where S and V are at zero and given as follows,

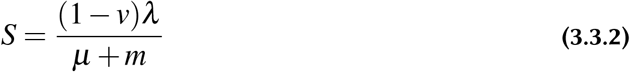

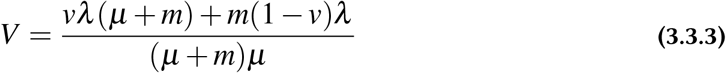

The other equations remain zero since all the compartments are equal to zero. Hence the disease-free equilibrium points are as follows:

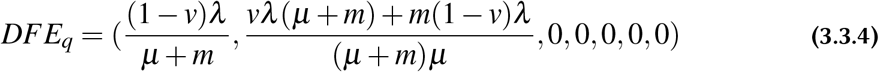

### 3.4 Reproduction number, Ro Computation

Reproduction number, *R*_0_, is the number expected of secondary infections of any infectious disease produced by a single or typical infection in a completely susceptible population [20]. Ro is a very important aspect since it plays a role in determining or measuring whether the infection will spread or not. In any given community or population, when *R*_0_ < 1, an infected individual will not meet the threshold of infecting a large number by just causing less than one new infection; hence there will be no spread of the virus which could make it die out. If *R*_0_=1, then each existing infectious individual will cause a new infection; thus, the infection will be stable and exist but will not spread to the rate of causing an epidemic.

When *R*_0_ > 1, each infected individual can cause more than one infection; hence the disease will spread rapidly in the population, persist and cause an epidemic.

Therefore, the basic reproduction number is key in making key decisions on COVID-19. If *x*_*o*_ is a DFE of *x*_*i*_ = *f*_*i*_(*x*) −*v*_*i*_(*x*) for i=1,2…n and *f*_*i*_(*x*) satisfies conditions:

1. If x≥0 then 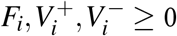 for i=1,2,. n.
2. If *x*_*i*_=0 then 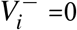
3. *F*_*i*_=0 if i>m.
4. If *x* ≥ **X**_*s*_ then *F*_*i*_(*x*)=0 and 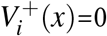 for i=1,…,m.
5. If F(x) is set to zero, then all the eigenvalues of Df(*x*_0_) have negative real parts.

Then the derivatives DF(*x*_0_) and DV(*x*_0_) are partitioned as

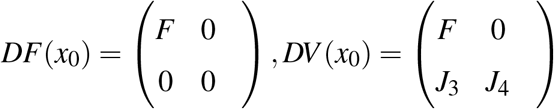

where F and V are m x m matrices defined by

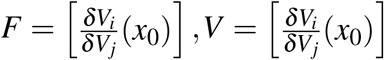

with 1≥i, *j* ≤m [22].

The next generation matrix *FV*^−^1 and the spectral radius of the next generation matrix *R*_0_ = *ρ*(*FV*^−^1)

To compute Ro, therefore, I must note the new infections from other variables in the model. Let F denote the rate of appearance of new infections into the infected compartments and V the transfer in and out of the infected compartments.

From the model equations,

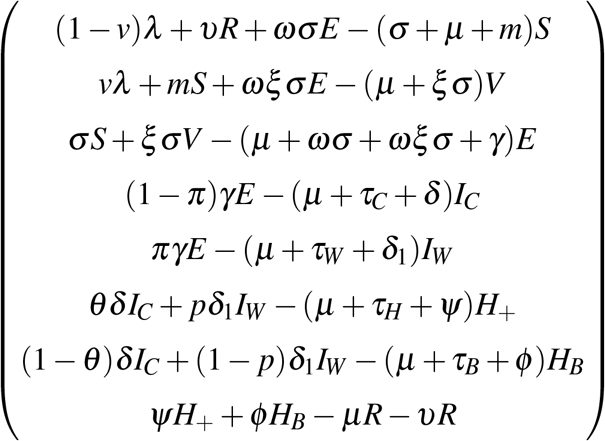

The rate at which the susceptible and vaccinated population get exposed is given as

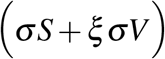

From the six categories between Exposed to Homebased care,I find,

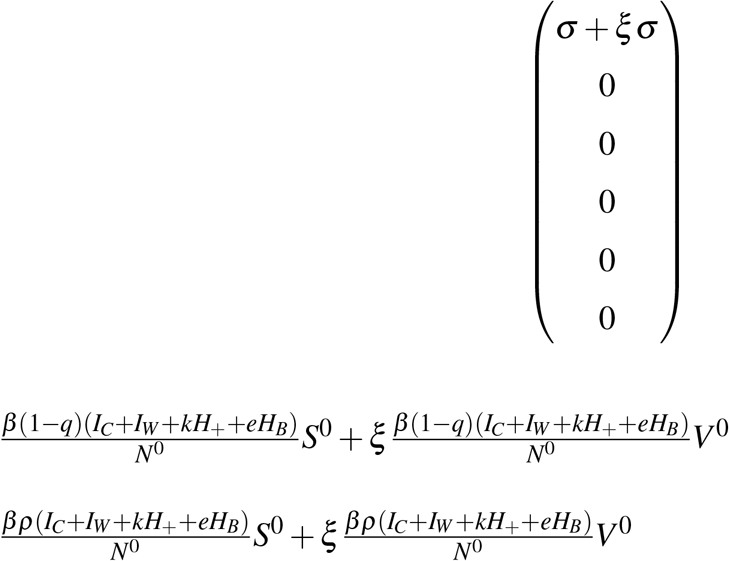

Therefore, F(y) becomes

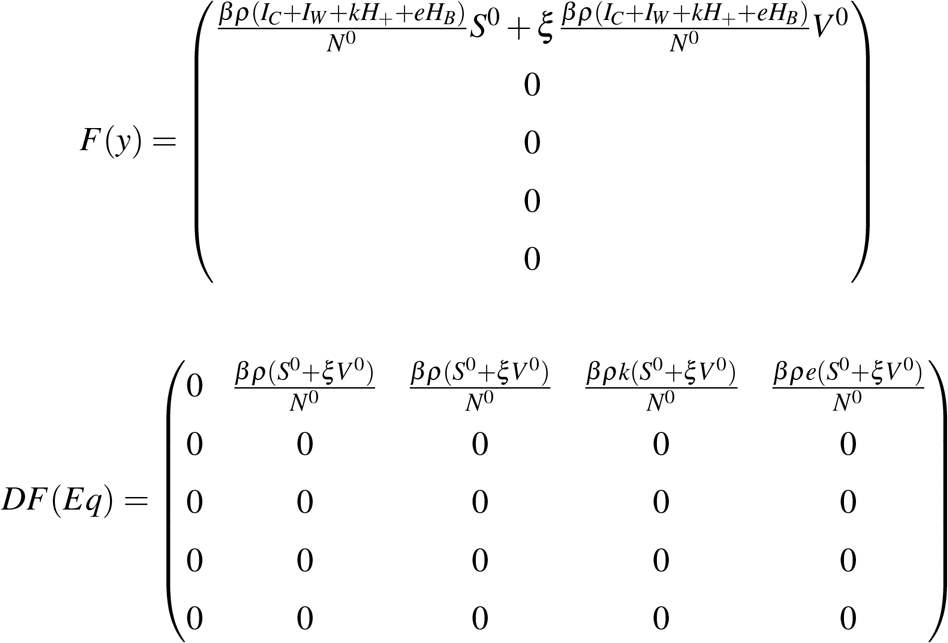

The disease free we have 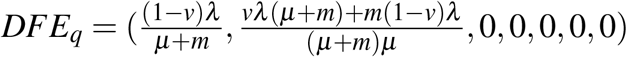

The rate of transfer of individuals in and out of the Infection compartment is given by

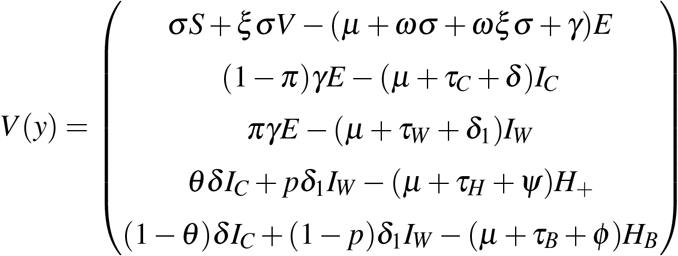

(Since the Susceptible and Vaccinated are not within the zone of infection or around the infection)

Rearranging V(y) to have

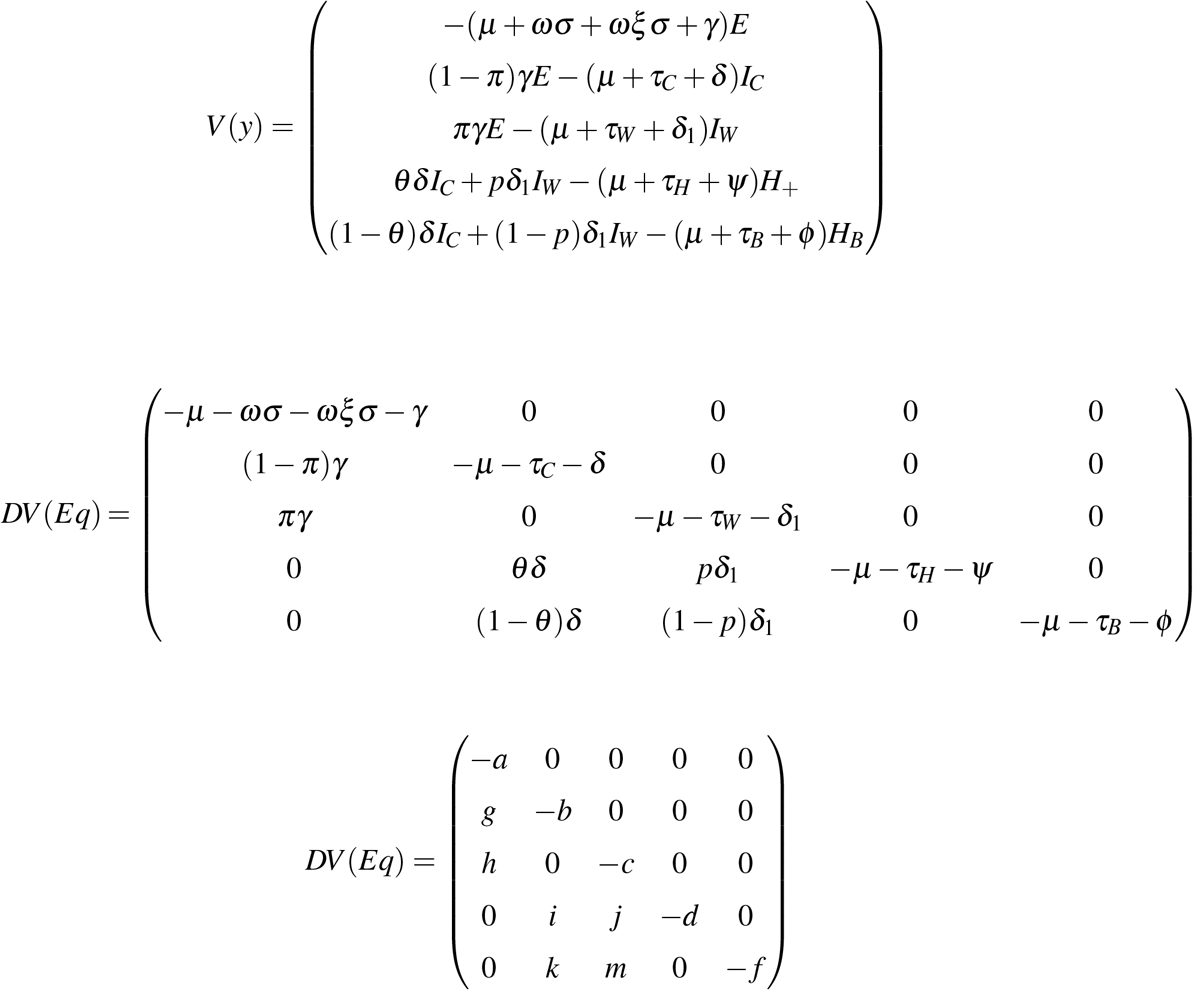

where a,b,c,d, f, g,h,i,j,k and m are *µ* + *ωσ* + *ωξσ* + *γ, µ* + *τ*_*C*_ + *δ, µ* + *τ*_*W*_ + *δ*_1_, *µ* + *τ*_*H*_ +*ψ, µ* + *τ*_*B*_ + *ϕ*, (1 −*π*)*γ, πγ, θδ, pδ*_1_, (1 −*θ*)*δ* and (1 − *p*)*δ*_1_

From here,

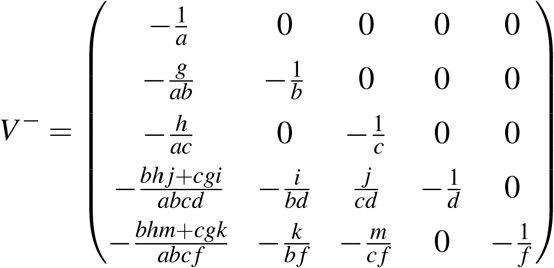

Multipliying F by *V*^−^ to get,

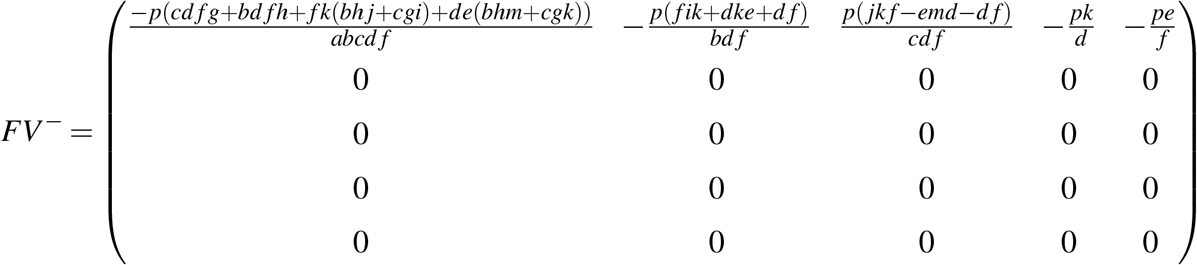

where 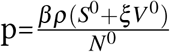

Since F is non-negative and V is non-singular, then *V*^−^ and *FV*^−^ are non-negative. *FV*^−^is the next-generation matrix for this model.

*R*_0_ is given by the spectral radius of the matrix D, which is *FV*^−^.

Since, *R*_*O*_=D,

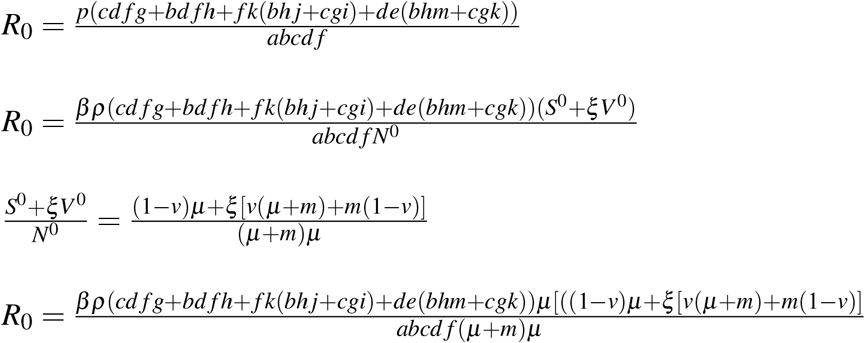

*R*_0_ is given as 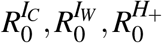 and 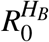

Here, 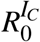 is the basic reproduction number of the proportion that are Infected and have other chronic illnesses,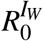 is the basic reproduction number of the proportion that are Infected and do not have other chronic illnesses,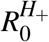 is the basic reproduction number of the proportion that are hospitalized and 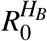 is the proportion that are under Home Based Care.

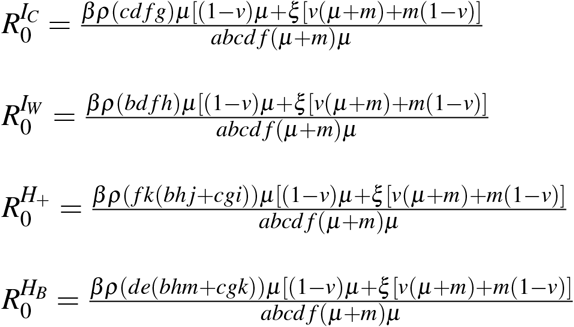

where a,b,c,d, f, g,h,i,j,k and m are *µ* + *ωσ* + *ωξσ* + *γ, µ* + *τ*_*C*_ + *δ, µ* + *τ*_*W*_ + *δ*_1_, *µ* + *τ*_*H*_ +*ψ, µ* + *τ*_*B*_ + *ϕ*, (1 −*π*)*γ, πγ, θδ, pδ*_1_, (1 −*θ*)*δ* and (1 − *p*)*δ*_1_

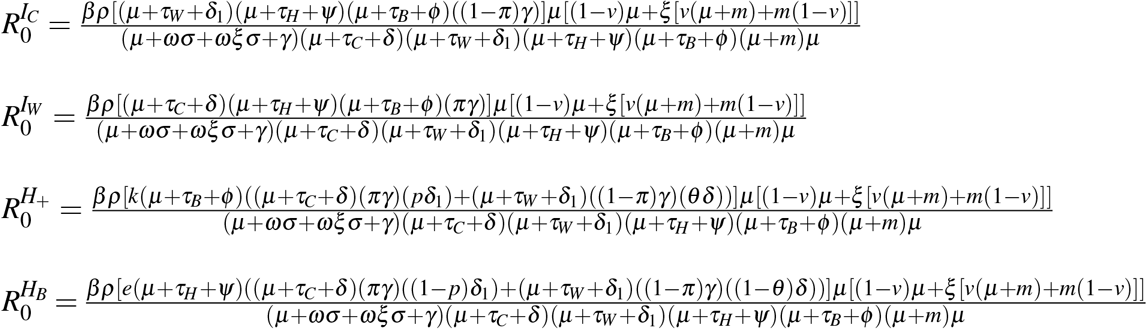

### 3.5 Stability Analysis

From my model,when there is no disease(COVID-19), the Exposed,Infected,Hospitalized,Home based care and Recovered become zero and the susceptible becomes 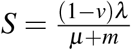 and the vaccinated 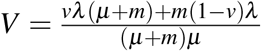therefore giving the equilibrium as,

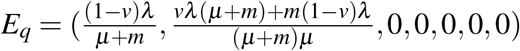

In this section,we are going to give a critical analysis of the local disease-free equilibrium, global disease-free equilibrium, local endemic equilibrium, and global endemic equilibrium.

#### Local DFE

The disease-free equilibrium, *E*_*q*_ of the model, is locally asymptotically stable if and only if *R*_0_< 1. It becomes locally unstable if *R*_0_> 1.

This means that in the Kenyan population, a patient (Patient zero) and other patients were introduced to the system. Some had other chronic illnesses, while others had no chronic illnesses. So in the event, the proceeding infections have an *R*_0_<1, then there will be no COVID-19 outbreak; on the other hand, if the proceeding infections have *R*_0_>1, then there will be an outbreak.

To analyse this, the system S(t), V(t), E(t), *I*_*C*_(*t*), *I*_*W*_ (*t*), *H*_+_(*t*), *H*_*B*_(*t*), R(t)

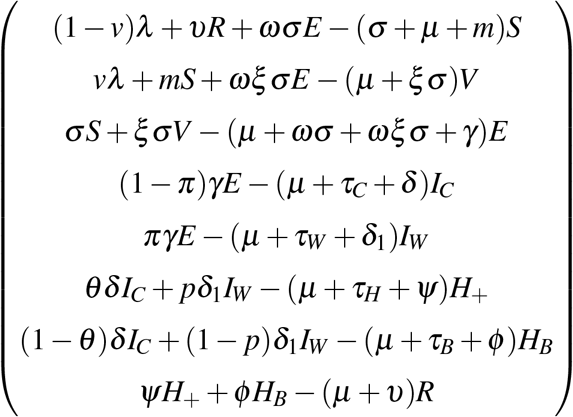

is linearized at the equilibrium point, bearing in mind that 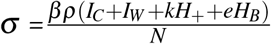, to give a Jacobian matrix in the form

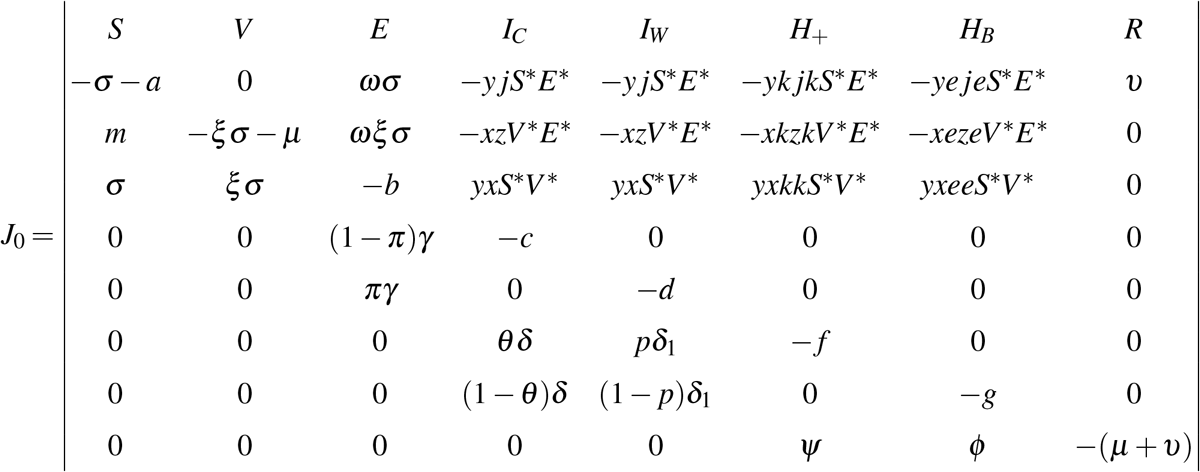

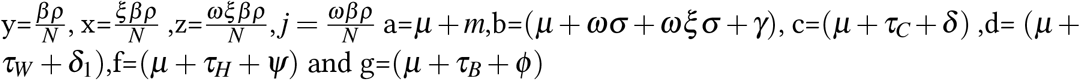

At the equilibrium point, the jacobian matrix becomes,

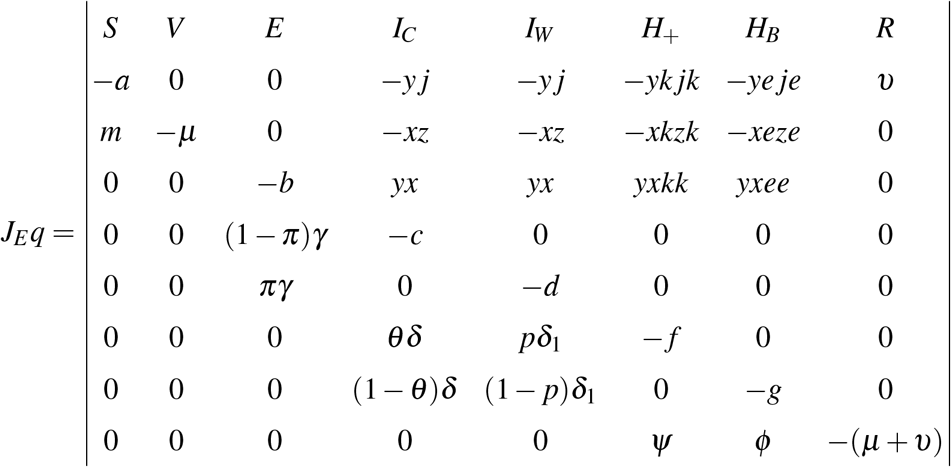

The eigen values of *J*_*E*_*q*

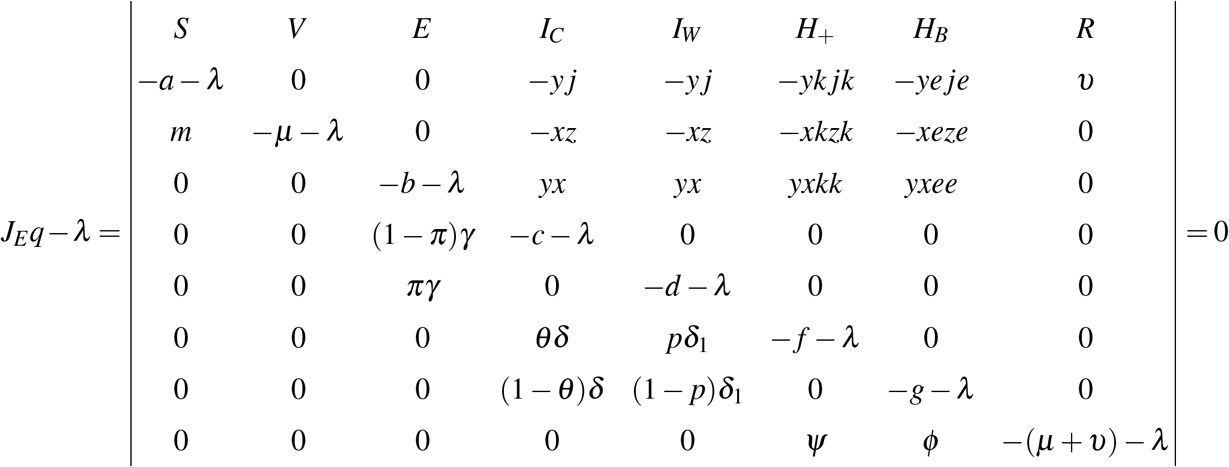

Solving this to yield the eigen values for the system as follows:

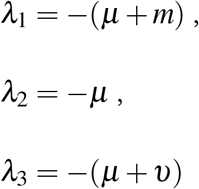

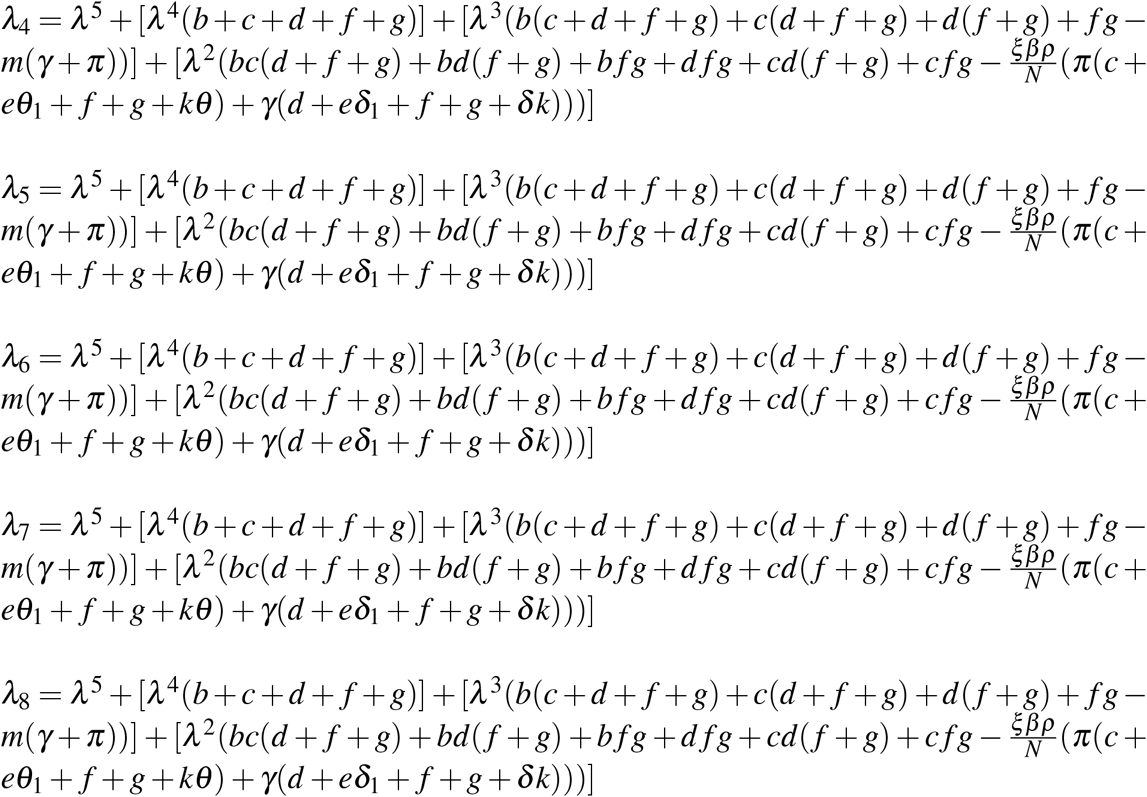

From these, the eigen values *λ*_1_, *λ*_2_ and *λ*_3_ are negative with positive parameters *µ, µ* + *υ* and *µ* + *m*

The eigenvalues show that the disease-free equilibrium of this model is asymptotically stable if *R*_0_<1.

#### Global DFE

Theorem: The disease-free equilibrium of this model is globally asymptotic stable if *R*_0_<1 and unstable if *R*_0_>1.

Proof: To establish the global disease-free equilibrium, a Lyapunov function is constructed:

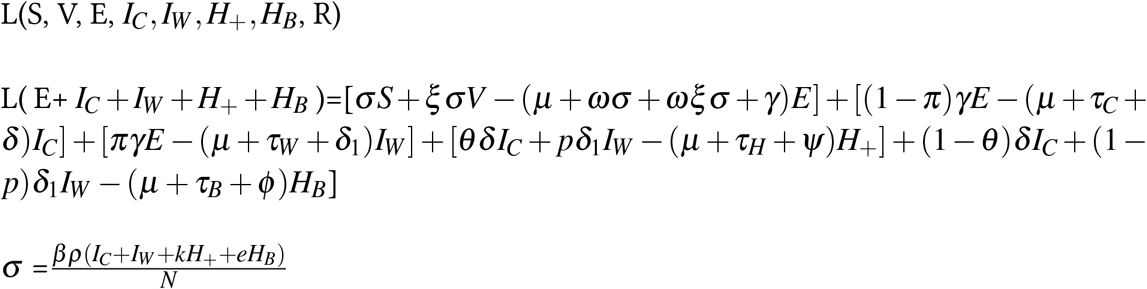

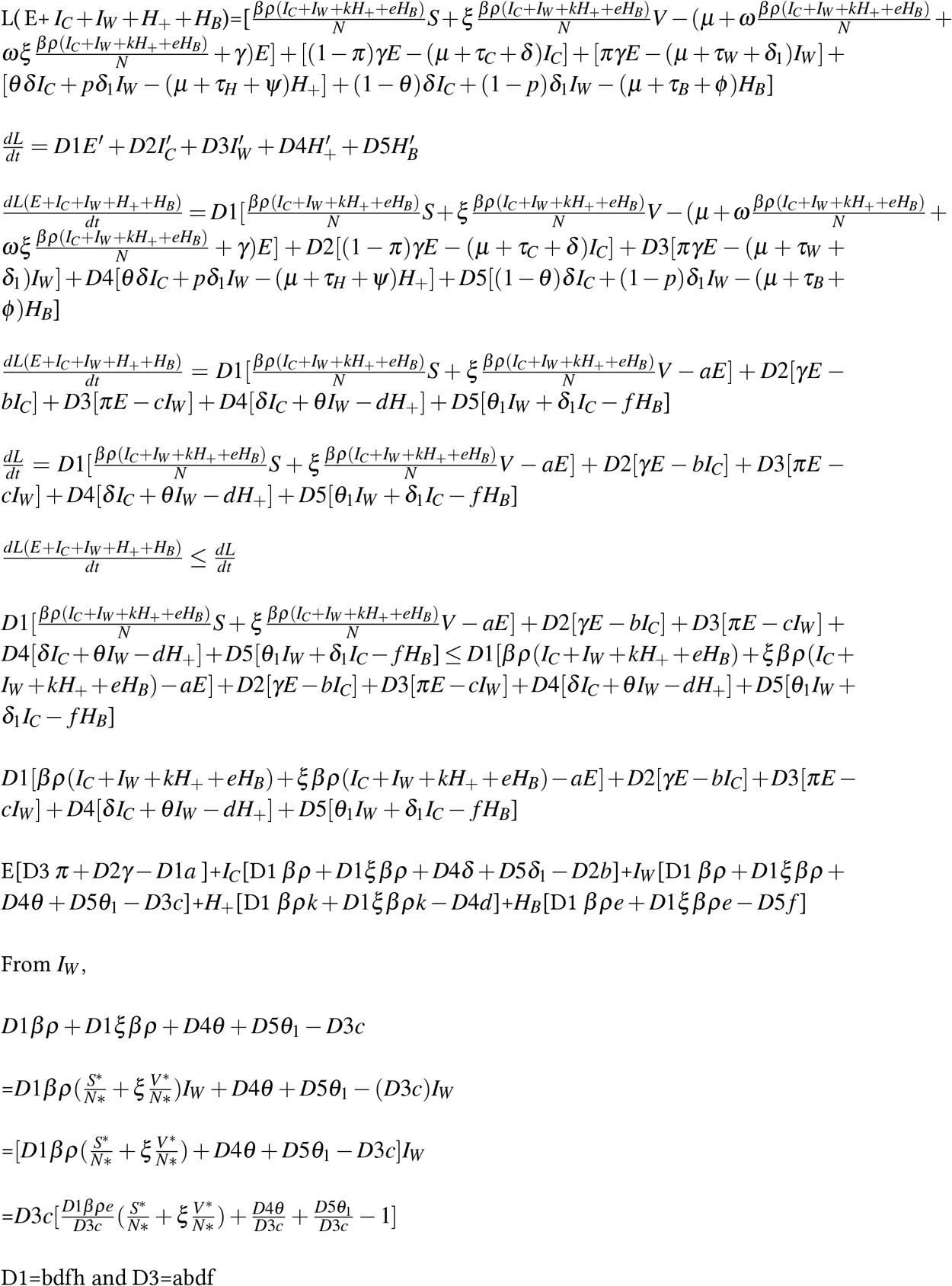

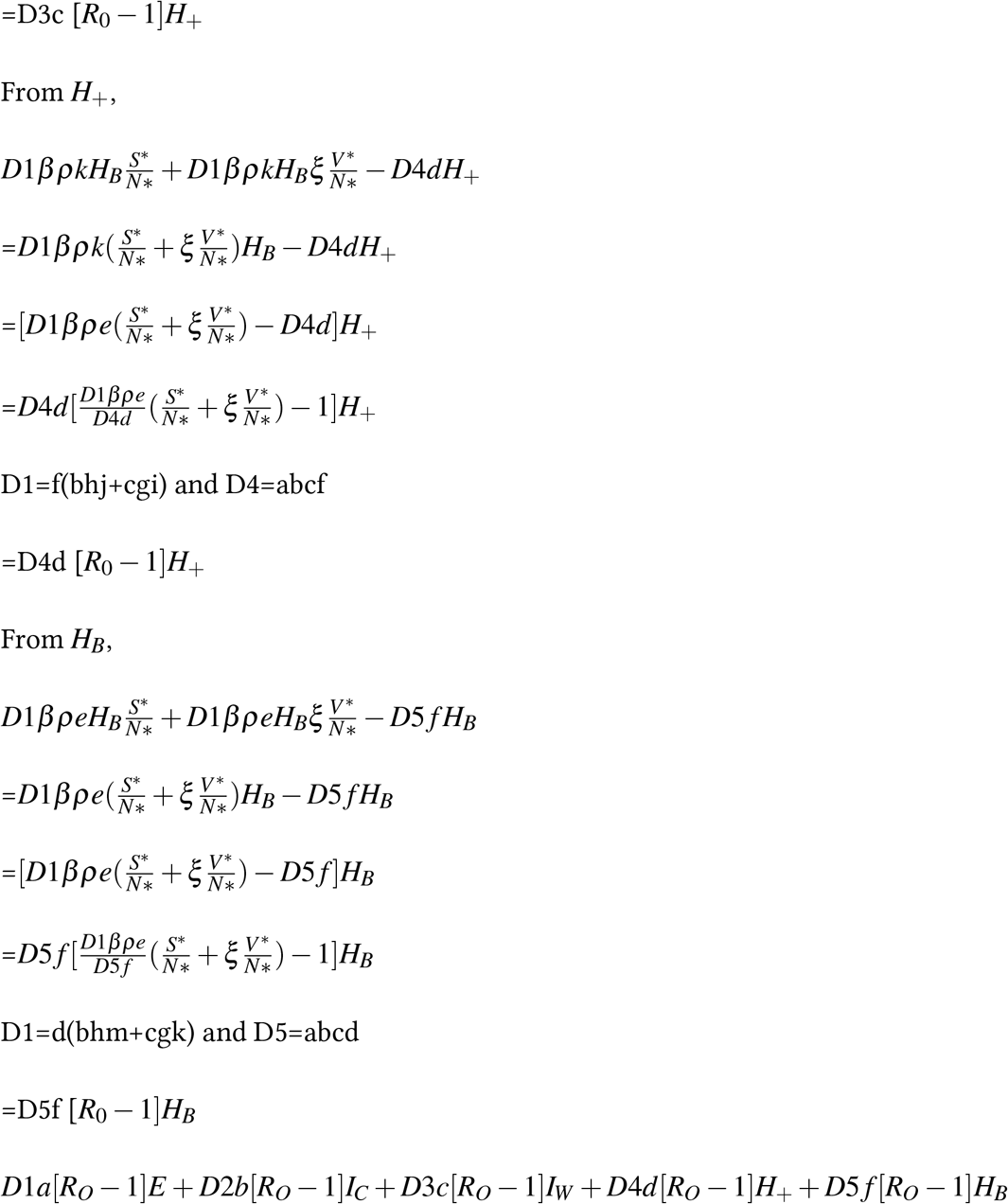

This means that 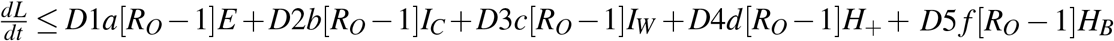

By the invariance theorem, the disease free equilibrium is stable when *R*_*O*_ ≥ 1.

### 3.6 Endemic Equilibrium

The endemic equilibrium point is a steady-state solution point where COVID-19 virus infections are persistent within the population.

It exists only and only if the number of susceptible is less than the total population.

The endemic equilibrium of the system is given by E=*S,V, E, I*_*W*_, *I*_*C*_, *H*_+_, *H*_*B*_, *R*. The model is set to zero and the force of infection 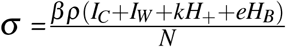

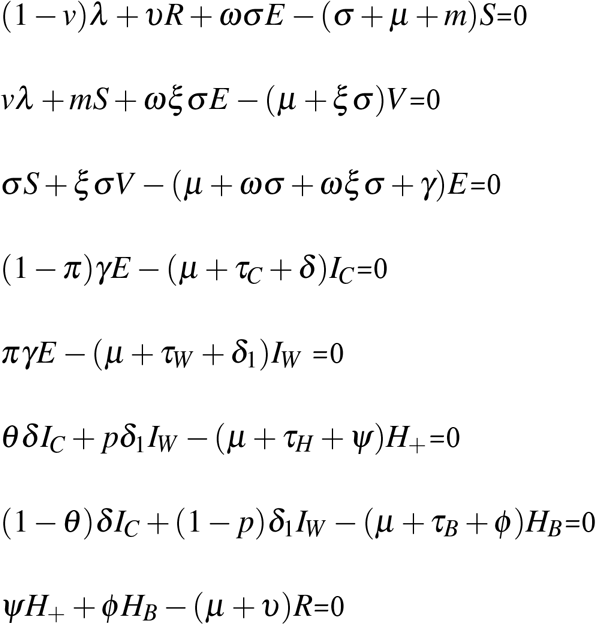

to obtain

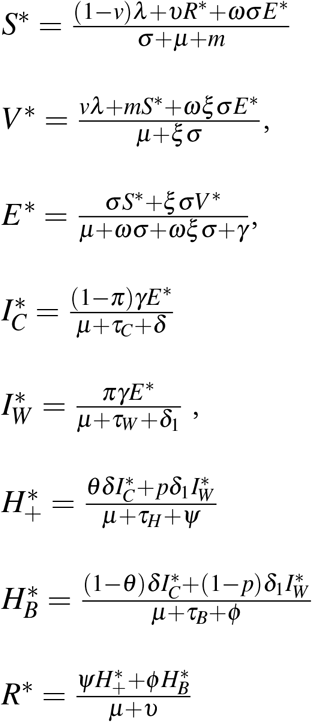

From this, let A=*σ* + *µ* + *m*, B=*µ* + *ξσ*, C=*µ* + *ωσ* + *ωξσ* + *γ*, D=*µ* + *τ*_*C*_ + *δ*, F=*µ* +*τ*_*W*_ + *δ*_1_, G=*µ* + *τ*_*H*_ + *ψ*, H=*µ* + *τ*_*B*_ + *ψ* and J=*µ* + *υ*

Therefore,

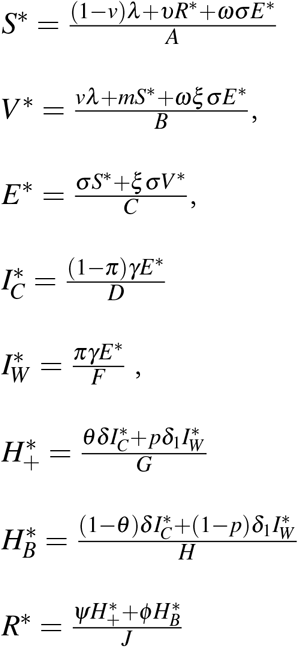

This finaly gives,

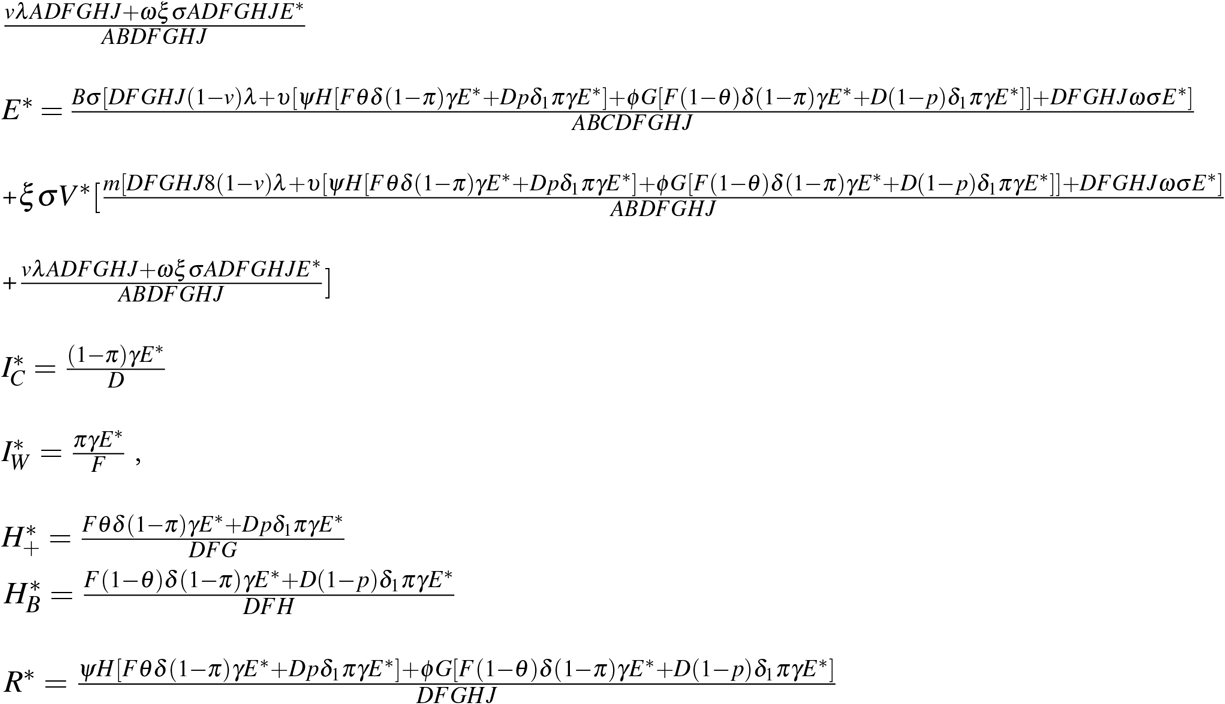

Here,

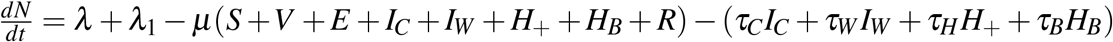

But, S+V+E+*I*_*C*_+*I*_*W*_ +*H*_+_+*H*_*B*_+R=N

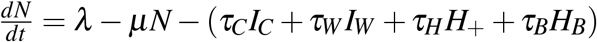

So 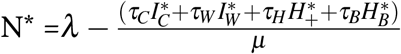

#### Local EE

The endemic equilibrium point, *E*_*q*_, of the model becomes asymptotically stable if and only if *R*_*O*_ >1.

To prove the stability of endemic equilibrium, the model equations are transformed into a Jacobian matrix hence.

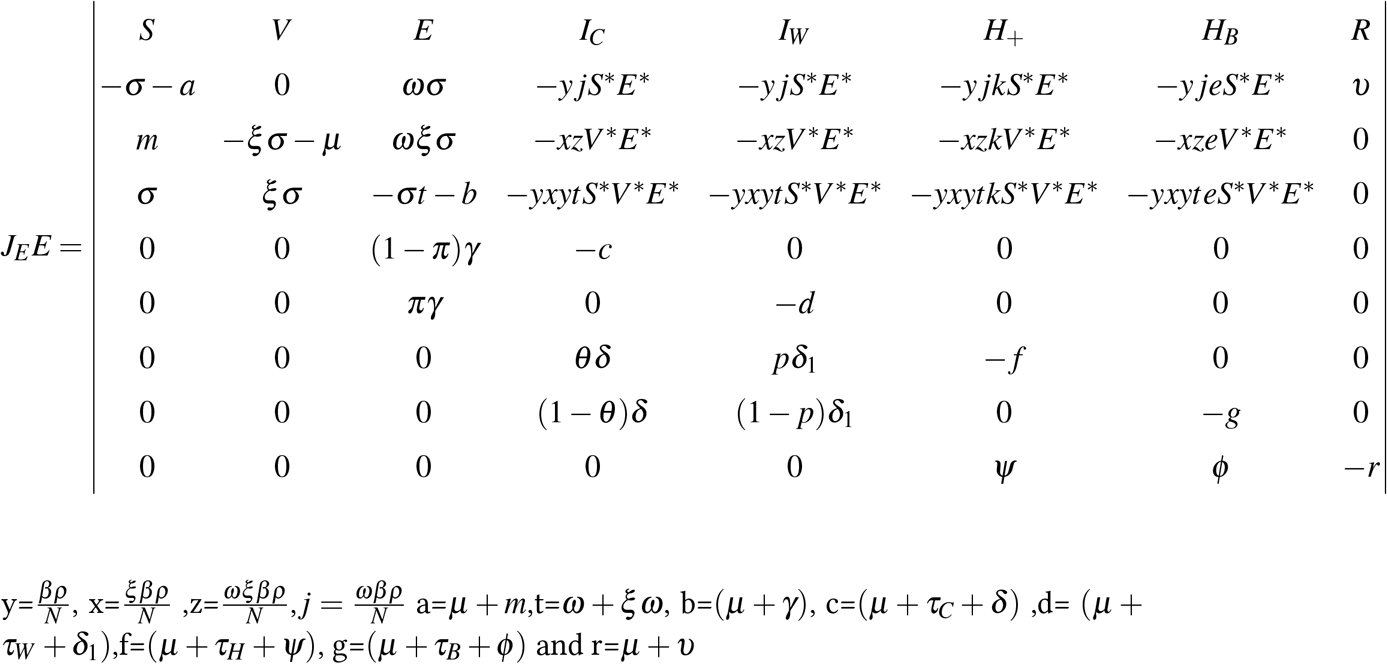

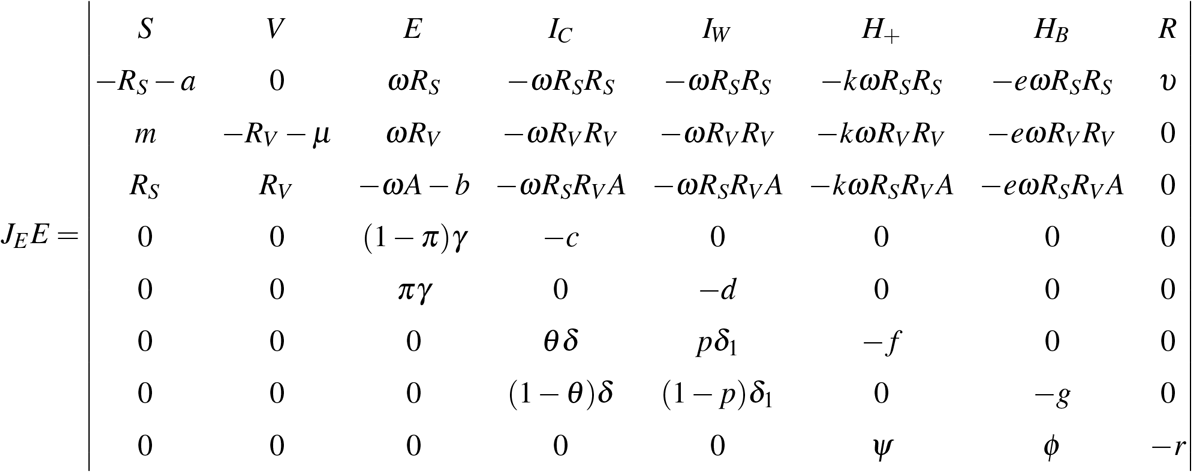

where A=*R*_*S*_ + *R*_*V*_

The eigen values of *J*_*E*_*E* is simplified as

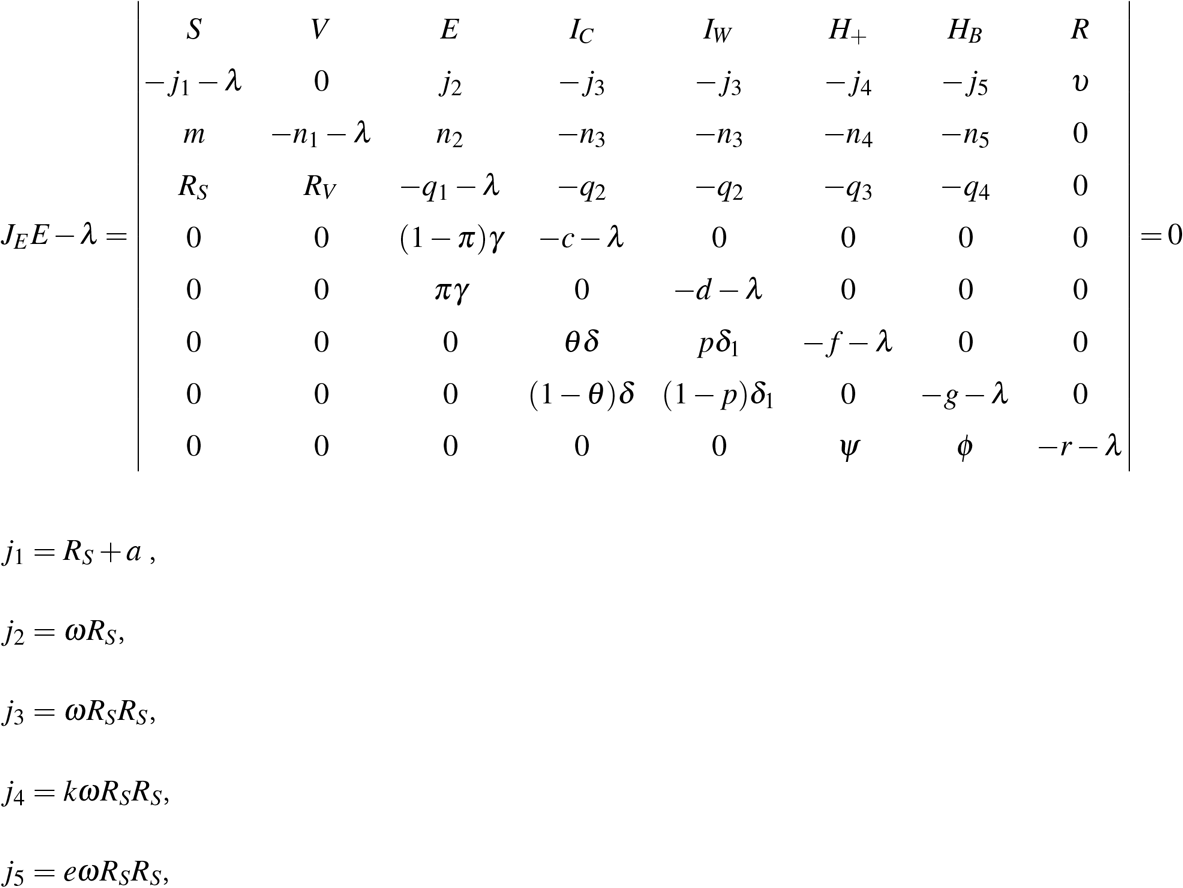

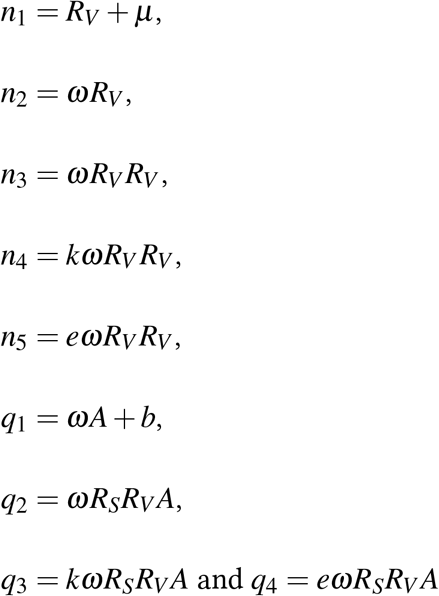

This takes the form 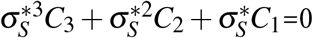

Where

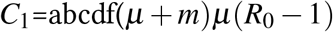

The system has a positive solution if *C*_1_ >1. Therefore, the model has a unique endemic equilibrium E^−^ if *C*_1_ >1.

#### Global EE

To analyze the global stability of the endemic equilibrium, we construct a Lyapunov function to define L(S, V, E, *I*_*C*_, *I*_*W*_, *H*_+_, *H*_*B*_, R):

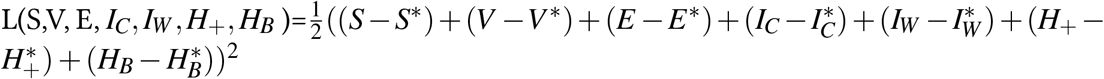

The Lyapunov function, L, is greater than zero. At the endemic equilibrium point, the function becomes equal to zero.

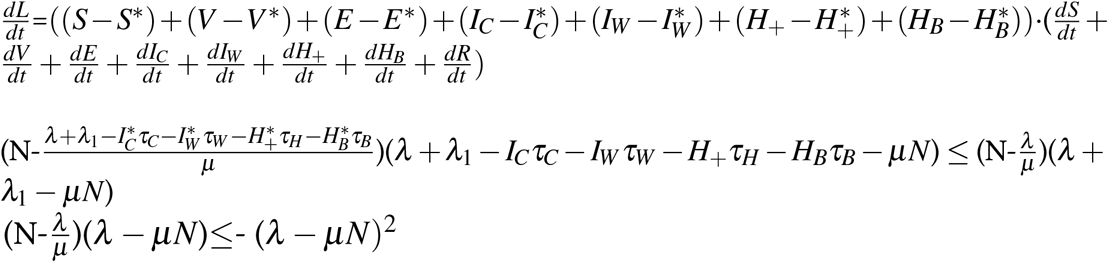

From this, it is evident that - (*λ* − *µN*)^2^ is ≤ 0 therefore,this invariant set,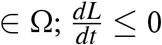 is the endemic equilibrium for the model

Thus by the LaSalle’s Invariance Principle (La Salle, 1976), S is globally asymptotically stable in Ω for *R*_0_ > 1.

## 4 Parameter Estimation, Numerical Simulations And Results

### 4.1 Parameter Estimation

The model parameters used in this study were extracted from other research work, and others were assumed or estimated as listed in Table 4. The assumed values are realistic.

These parameters were taken from research done by Scholars in Modeling the Virus using Kenyan data. These resources or previous works are Mathematical Models of COVID-19 Transmission in Kenya:A Model with Reinfection Transmission Mechanism, Computational and Mathematical Methods in Medicine,[14], S-E-I-R model for COVID-19 dynamics incorporating the environment and social distancing.[21], Age-structured model for COVID-19: Effectiveness of social distancing and contact reduction in Kenya.[24], COVID-19 outbreak, social distancing and mass testing in Kenya-insights from a mathematical model.[25] and the Kenyan Population according to the Kenya Population and Housing Census 2019 report from Kenya National Bureau of Statistics, KNBS.

The assumed parameters were also sought from a realistic range of variables from previous research.

Table 4 shows the Parameters, values, and sources.

From this table, the sign of the values of these parameters means the degree of influence they have in spreading the disease. The positive indices mean that they have a positive impact on spreading the virus. In contrast, the negative indices mean they have no impact in causing the spread of the virus—the basic reproduction number changes when the seven positive parameters are changed.

Parameters *β, ρ, ξ, k, e, m* and *π* have positive values and this means that they play a key role is in the spread of COVID-19 virus.When these parameters are changed, the basic reproduction number also changes

### 4.2 Numerical Simulations

Numerical simulations were carried out on the model system in Figure 1 to test COVID 19 behaviour. MoH and WHO aim at reducing infections to fully supress the virus.

**Table 1.** Parameters, used values, and their sources.

| Number | Parameter | Value | Source |
| --- | --- | --- | --- |
| 1 | $\omega$ | 0.0051 | [21] |
| 2 | $\tau_C$ | 0.0020 | [21] |
| 3 | $\tau_W$ | 0.0016 | [21] |
| 4 | $\tau_H$ | 0.0017 | [21] |
| 5 | $\tau_B$ | 0.0019 | [21] |
| 6 | $\delta$ | 0.35 | [14] |
| 7 | $\delta_1$ | 0.19 | [14] |
| 8 | $\psi$ | 0.064 | [24] |
| 9 | $\phi$ | 0.042 | [25] |
| 10 | N | 47,564,296 | [26] |
| 11 | $\lambda$ | 3000 | Assumed |
| 12 | v | 0.38 | Assumed |
| 13 | $\nu$ | 0.0078 | Estimated |
| 14 | $\xi$ | 0.5 | Assumed |
| 15 | q | 0.4 | Estimated |
| 16 | $\beta$ | 0.56 | Estimated |
| 17 | $\gamma$ | 0.189 | Assumed |
| 18 | $\pi$ | 0.4 | Assumed |
| 19 | $\theta$ | 0.65 | Estimated |
| 20 | p | 0.4 | Assumed |
| 21 | $\mu$ | $4.076 \times 10^{-5}$ | Estimated |
| 22 | k | 0.62 | Assumed |
| 23 | e | 0.45 | Assumed |
| 24 | m | 0.1166 | Estimated |

We conducted various simulations using various control measures ranging from compliance or afdherence to MoH /WHO health standards, vaccination rates, vaccine effectiveness,reinfection rates and infection reduction due to hospitalization and home based care.

These simulations were done using MATLAB

#### Model Simulation

The simulation for the eight compartments without control was done using the equations in Figure 1 and parameters in Table 4 to show the model’s eight compartments. These simulations were done when V=12,N=47.6 Million,E=600,*I*_*C*_ = 740, *I*_*W*_ = 459, *H*_+_ = 20, *H*_*B*_=33 and R=3.

##### Varying rates of compliance to the measures

Compliance rates to the MoH guidelines were varied to show the rates of the Exposed, Infected, and Hospitalized. The variations were done for p=0.8,0.6,0.4 and 0.2.

##### Varying vaccination rate

The variations were done for m=0.01,0.03,0.05 and 0.07

##### Varying vaccine effectiveness

The variations were done for vaccine effectiveness at 50 percent, 80 percent, 90 percent and 100 percent.

##### Varying reinfection rates

The reinfection rates, *υ* were varried at diffrent rates of 0.008,0.010,0.012 and 0.014

### 4.3 Results

In this section, we are going to give a brief explanation of the simulations with respect to different control strategies.

For the basic reproduction number,

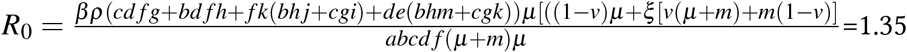. When there is no vaccination,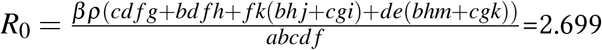

#### 4.3.1 Simulation with no Strategy

**Figure 1.**
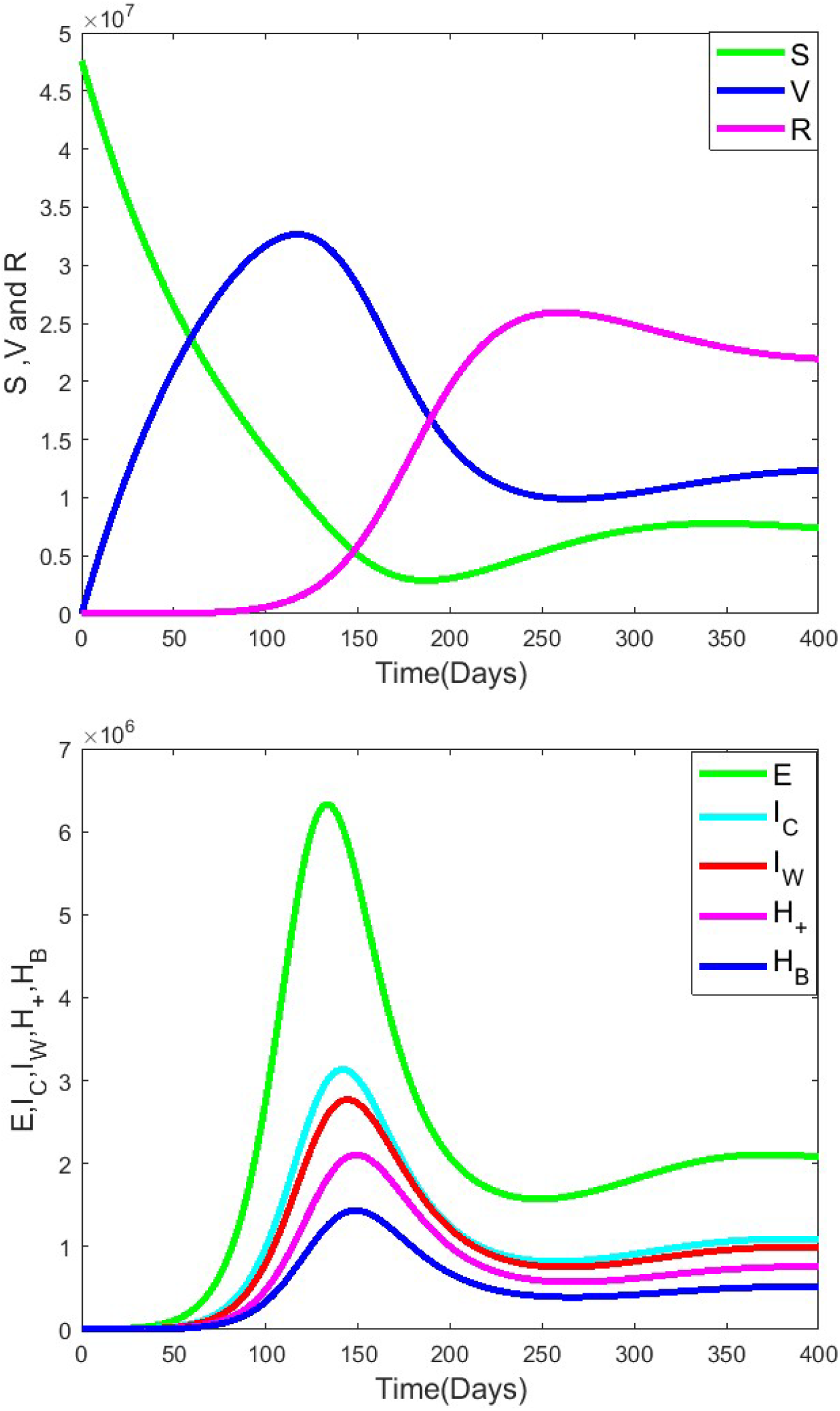
The eight compartments.

**Figure 2.**
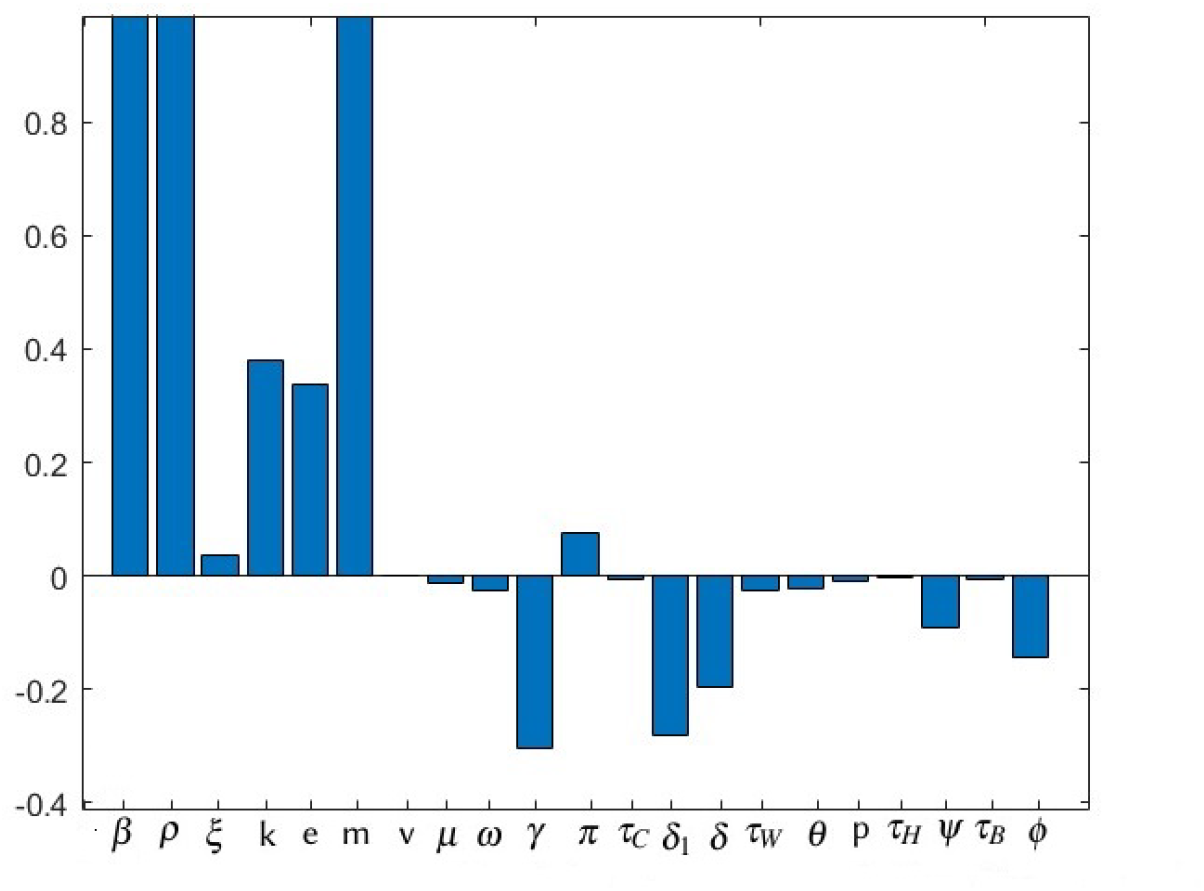
Sensitivity Indices of *R*_0_ Parameters.

**Figure 3.**
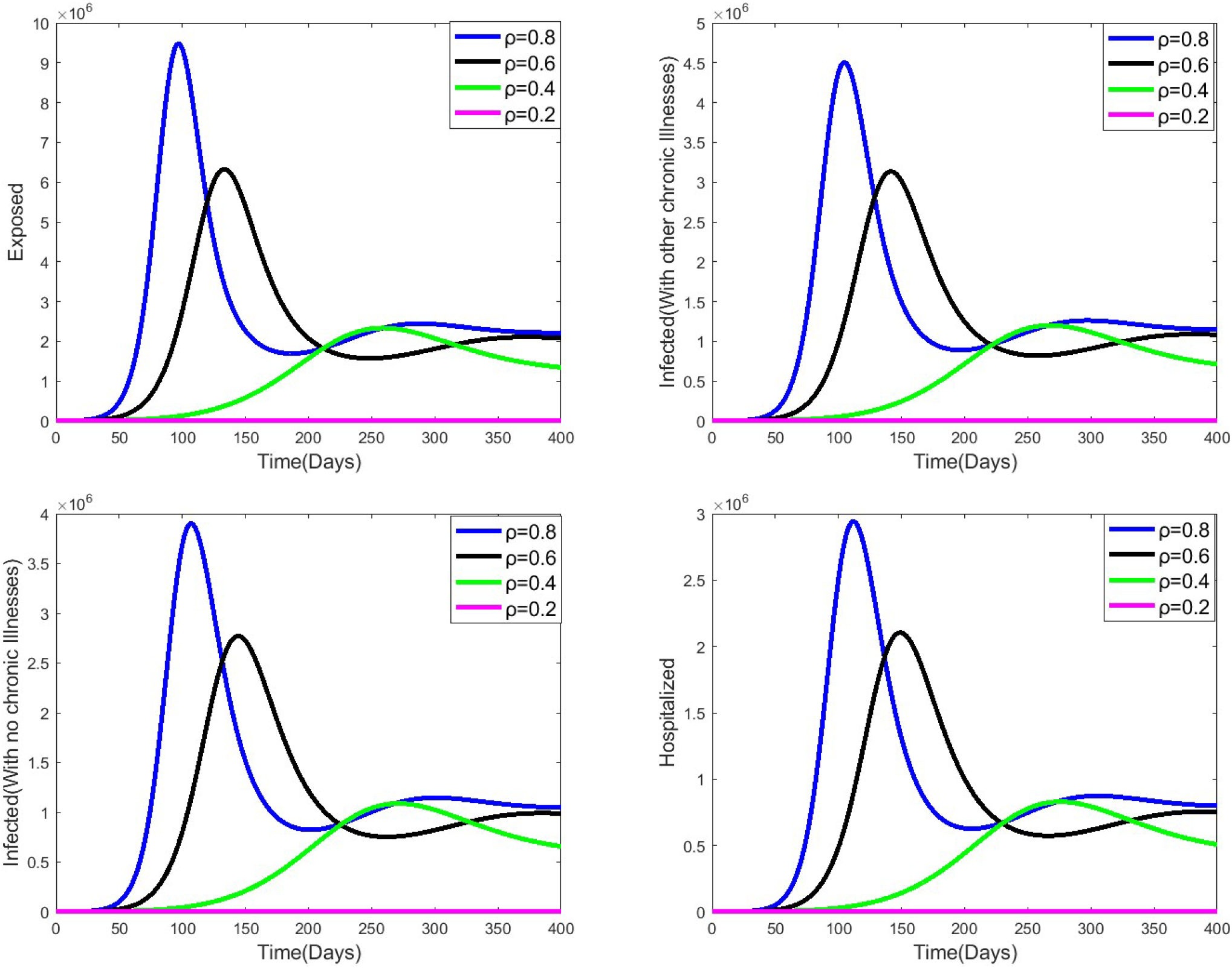
Compairing Exposure,Infection and Hospitalization when *ρ* is varried.

**Figure 4.**
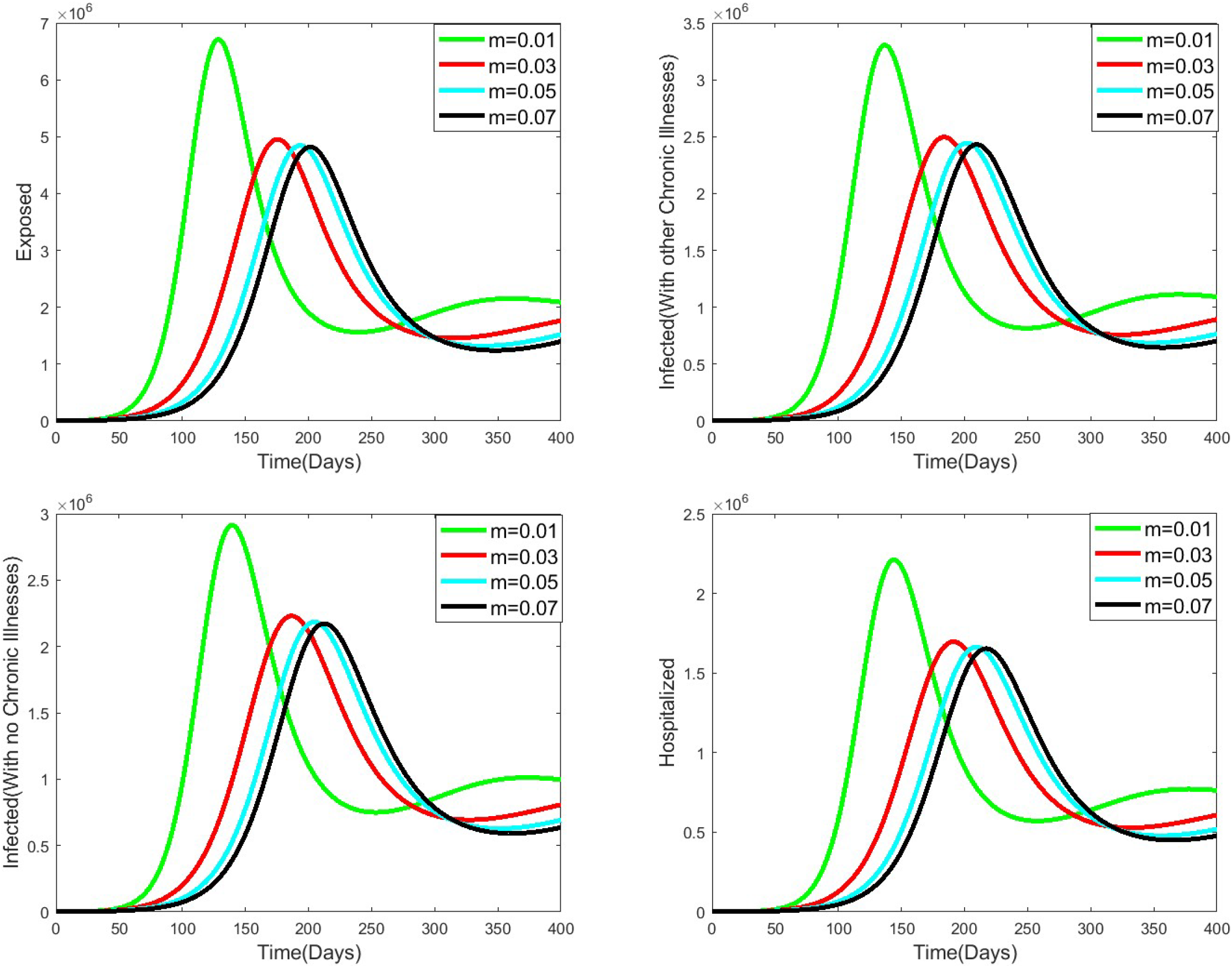
Compairing Exposure,Infection and Hospitalization when m is varried.

**Figure 5.**
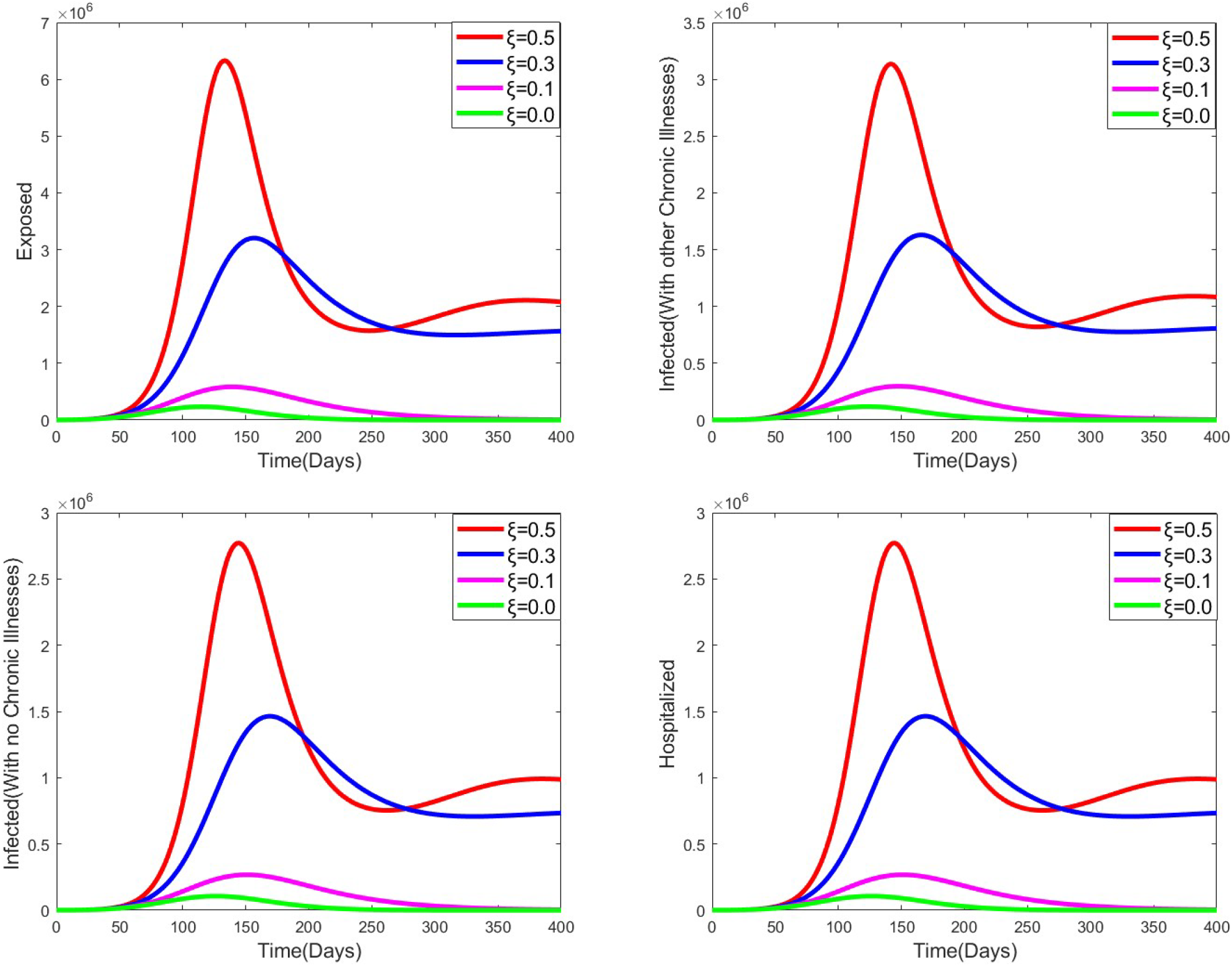
Compairing Exposure,Infection and Hospitalization when effectiveness of the vaccine is varried.

**Figure 6.**
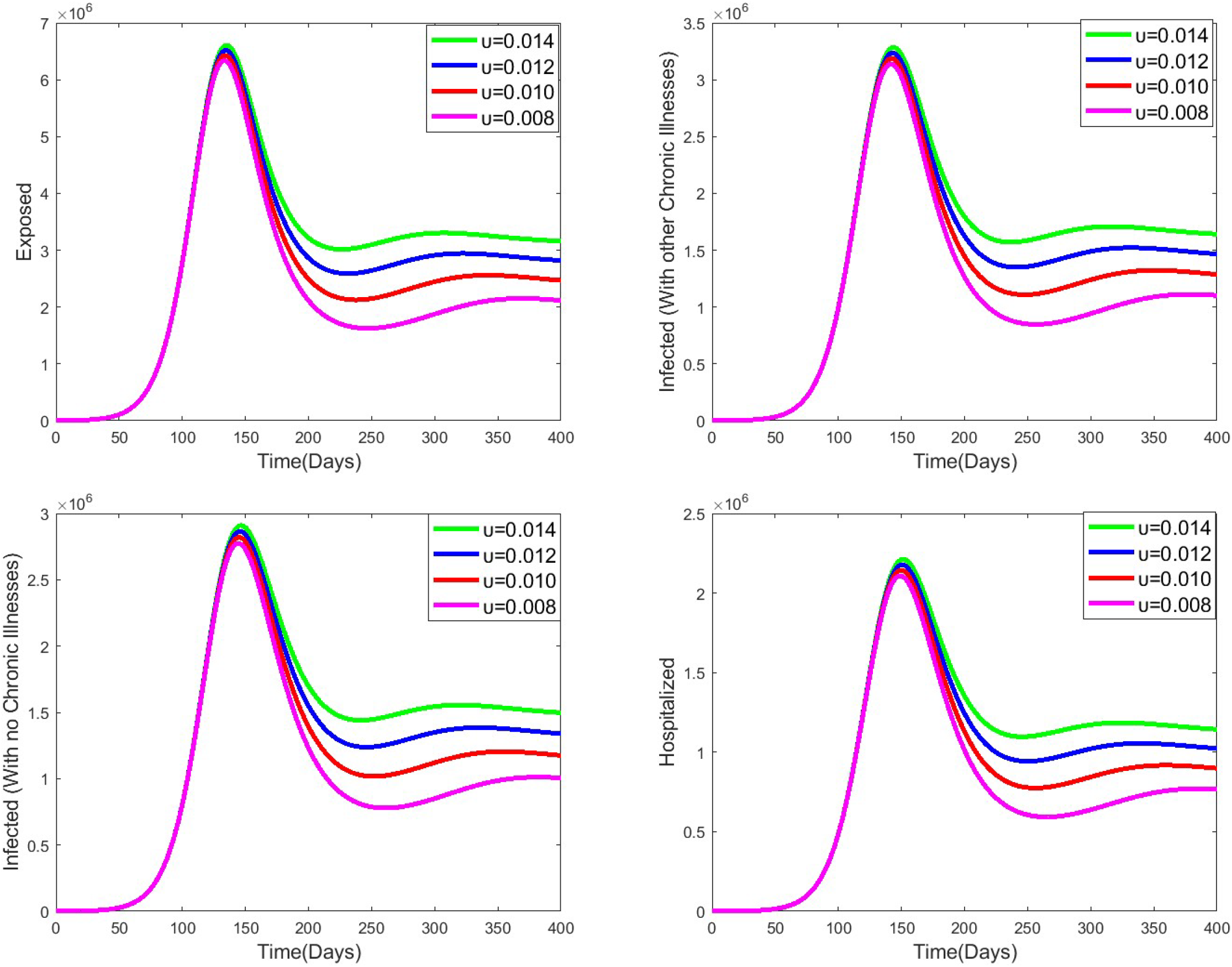
Compairing Exposure,Infection and Hospitalization when reinfection, *υ* is varried.

**Figure 7.**
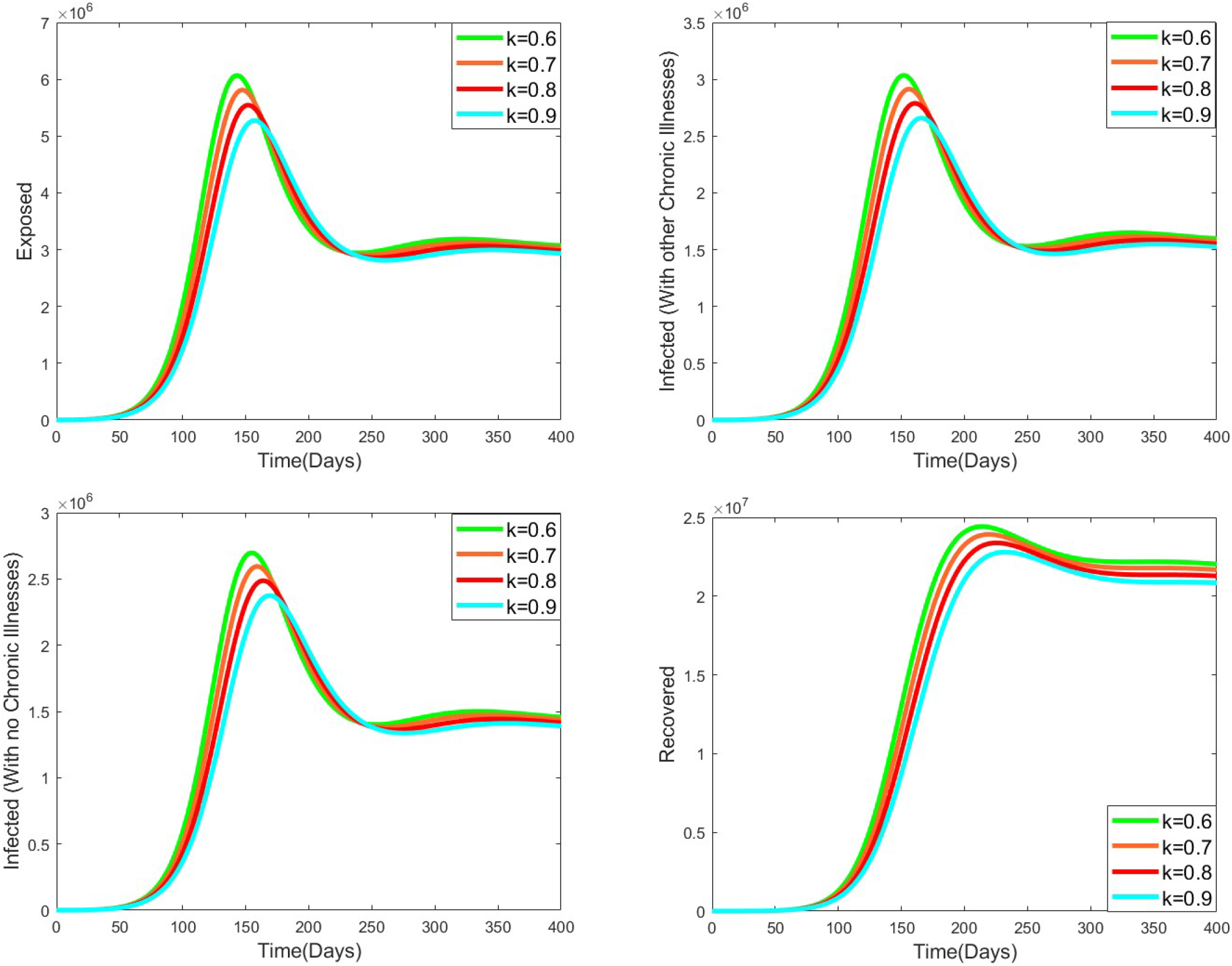
Compairing Exposure,Infection and Recovery when infection reduction due to hospitalization,k is varried.

**Figure 8.**
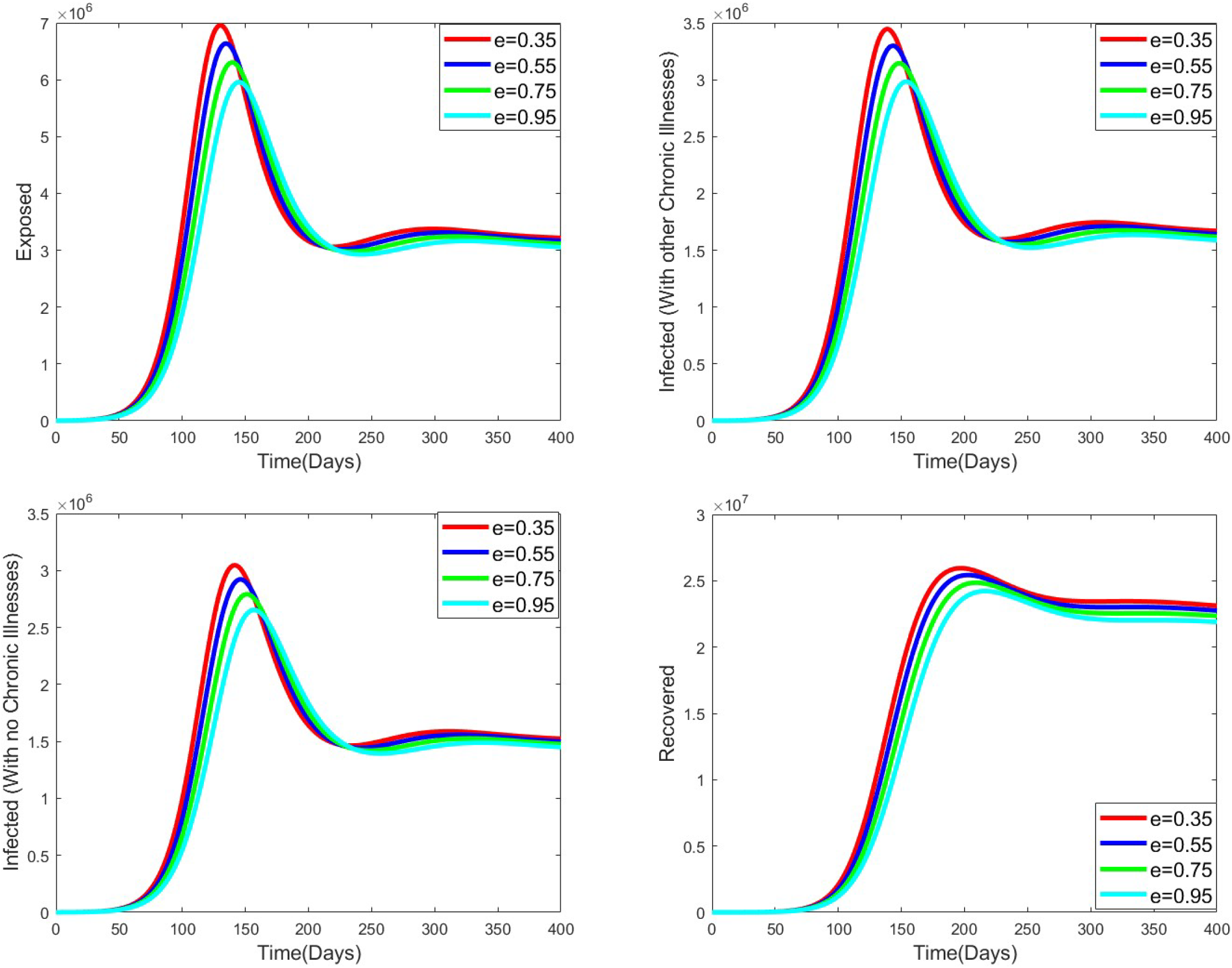
Compairing Exposure,Infection and Recovery when infection reduction due to home based care,e is varried.

The simulation shows a steady rise in Exposure to the 150 days, this leads to subsequent infections, hospitalization, and home-based care. The Susceptible proportion lowers to day 170 and then rises at a slow pace.

#### 4.3.2 Varying rates of compliance to the measures

In this section, we varied the compliance rate. It shows that when the compliance rate is increased, the virus infection spread reduces considerably. When the compliance rate is 0.6, the Exposed peaks at 2.5 Million people after 270 days, whereas when it is lowered to 0.2, then the Exposed is 9.5 Million at 100 days.

#### 4.3.3 Varying vaccination rate

Vaccination plays an important role in the spread of COVID-19. When the vaccination rate is 0.01, the number of people who will be infected and have other chronic illnesses will be 3.5 million after 150 days, and when it is increased to 0.3, then only 2.5 will be infected. From this simulation, the curves for 0.03,0.05, and 0.07 are almost similar; this shows even if the vaccination coverage is high but vaccine effectiveness is low, the virus cannot be eradicated.

#### 4.3.4 Varying vaccine effectiveness

Vaccine effectiveness is key. Its variation has an impactful result. From our simulations, when it is 50 percent effective, then 6.5 million people will be exposed at maximum in 150 days; when it is increased to 80 percent effectiveness, the Exposure lowers to 3 Million, which is a big margin when compared to the 50 percent effectiveness. At 90 percent, 500,000 people will be exposed to the disease. This dictates that researchers should work to improve the effectiveness of the vaccine to lower the spread of the virus.

#### 4.3.5 Varying reinfection rates

The reinfection of previously infected individuals influences the spread of the virus. From the simulations, after 150 days, reinfection rates determine the new infection; when it is at 0.014, then infected people will be 1.5 million at day 250 and 0.7 million at day 250 when reinfection is lowered to 0.008. This means that sensitization should be done to the people who have recovered to still follow the set guidelines in curbing the spread of the virus.

#### 4.3.6 Varying infection reduction rate due to hospitalization

When an infectious individual is hospitalized, the chances of spreading the disease go down. At 0.9, the infections are at 2.3 million after 150 days, then flattens at day 250 going forward. When the infection reduction is 0.6, then new infections will peak on day 150, and they will be 2.7 million cases. This means that to increase infection reduction due to hospitalization. Health practitioners must be trained on how to handle COVID patients properly and equipped with enough personal protective equipment and PPE. The patients under care must also follow guidelines within the health facilities so as to reduce infection rates.

#### 4.3.7 Varying infection reduction rate due to home based care

The home-based care program that the Ministry of Health rolled out has also reduced new infections. When Infection reduction due to home-based care is at 0.35, the Exposed individuals will be 7 million, and when it is increased to 0.95, the Exposed will be 5.5 million, peaking after 150 days. This means that caregivers at home should be sensitized on how to handle COVID patients at home, and the patients should also adhere to the home-based care guidelines to lower the spread of this virus.

## 5 Conclusion

### 5.2 Conclusion

At the onset of the pandemic, medics determined that an individual could get reinfected, and others developed immunity against the virus. The introduction of COVID vaccine lowered the chances of being infected. Compliance with the MoH and WHO guidelines also dictated the infection rates and thus formed the basis of our thesis.

This deterministic model has been developed into eight compartments/proportions; the Susceptible, Vaccinated, Exposed, Infected(with other chronic illnesses), Infected(with no chronic illnesses), Hospitalized, Home based care, and Recovered to understand the infection and recovery dynamics of COVID-19 in Kenya. The people in the vaccinated compartment are considered to have been partially or fully vaccinated.Force of infection, 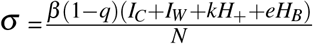 where compliance to set guidelines, infection reduction due to home-based care and hospitalization are considered within the Kenyan population.

We performed analysis to prove the positivity and boundedness of the model equations. The local and global disease-free and endemic equilibrium were done using the Jacobian Matrix and Lyapunov functions. The stability analysis of the model was conducted to determine those parameters that are key in driving the infections among the population from the basic reproduction number is 1.35 which means that an infectious individual can infect two people.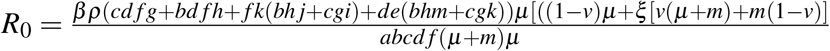

The study shows that compliance with the set guidelines imposed by the Ministry of Health to control the spread of COVID-19 plays a key role. When compliance is low, the basic reproduction number increases, meaning that the number of secondary infections from individuals increase. When the public keeps the guidelines, the infection rate within the population lowers, and it is evident that at 90 percent compliance, the virus will not be endemic because the basic reproduction number, *R*_*O*_, becomes less than 1.

Finally, we did the model system’s numerical simulation using MATLAB software. For the numerical simulation, we take the initial data and simulate using the parameters we got from other sources and others that are assumed. To reduce the spread of COVID 19 virus infection, we employed the controls like compliance to set guidelines, vaccination, hospitalization, and home-based care to minimize the objective function. The numerical results show that an increase in controls lowers COVID-19 spread.

## Data Availability

All data used are from public sources located in
M.O.H KENYA . Frequently Asked Questions on COVID-19 Vaccination 2021
JOHN HOPKINS UNIVERSITY COVID-19 Dashboard 31 March 2020
W.H.O..Weekly epidemiological update - 29 December 2020 2020

https://www.health.go.ke/covid-19

https://coronavirus.jhu.edu/about/how-to-use-our-data

https://www.who.int/publications/journals/weekly-epidemiological-record

